# AI-Enabled Diagnostic Prediction within Electronic Health Records to Enhance Biosurveillance and Early Outbreak Detection

**DOI:** 10.1101/2025.05.14.25327606

**Authors:** Andre R. Goncalves, Jose Cadena Pico, Yeping Hu, David Schlessinger, John Greene, Liam O’suilleabhain, Heather Clancy, Michael Vollmer, Vincent Liu, Tom Bates, Priyadip Ray

## Abstract

Detecting infectious disease outbreaks promptly is crucial for effective public health responses, minimizing transmission, and enabling critical interventions. This study introduces a method that integrates machine learning (ML)-based diagnostic predictions with traditional epidemiological surveillance to enhance biosurveillance systems. Using 4.5 million patient records from 2010 to 2022, ML models were trained to predict, within 24-hour intervals, the likelihood of patients being diagnosed with infectious or unspecified gastrointestinal, respiratory, or neurological diseases. High-confidence predictions were combined with final diagnoses and analyzed using spatiotemporal outbreak detection techniques. Among diseases with five or more outbreaks between 2014 and 2022, 33.3% (41 of 123 outbreaks) were detected earlier, with lead times ranging from 1 to 24 days and an average of 1.33 false positive outbreaks detected annually. This approach demonstrates the potential of integrating ML with conventional methods for faster outbreak detection, provided adequate disease-specific training data is available.

## Introduction

Rapid detection of population-based health anomalies is critical for enabling timely public health responses, including the implementation of potentially life-saving interventions and strategies to limit further transmission. Early identification of infectious disease outbreaks, such as those caused by respiratory or foodborne pathogens, is particularly crucial, as public health interventions can significantly mitigate their impact [1, 2]. Moreover, this capability supports biodefense missions by facilitating the rapid detection of intentional releases of hazardous substances, thereby enabling an accelerated response to minimize consequences [3].

Traditional public health surveillance relies on planning, organizing, analyzing, interpreting, and communicating surveillance information as the basis for public health decision-making [4]. However, the timeliness of infectious disease outbreak detection can be compromised due to its dependence on high-confidence diagnoses and, in some cases, laboratory confirmation before official communication to appropriate health authorities. Efforts to develop biosurveillance systems that improve the timeliness of health anomaly detection broadly fall into two categories: (1) the utilization of non-traditional data streams that may provide earlier indicators (syndromes or events) of an outbreak using typically non-clinical and non-laboratory-based data, and (2) the direct use of electronic health record (EHR)-based data, which is collected at scale during routine care.

Exploration of non-traditional data streams for biosurveillance has encompassed various sources, including ambulance dispatch data [5], school absenteeism records [6], Wikipedia queries [7], poison control center call logs [8], pharmaceutical sales [9], and many others. While these data streams are appealing due to cost savings from repurposing existing data, they often lack specificity and necessitate follow-up investigations before becoming actionable for public health officials. This additional step may diminish the benefit of early recognition of potential outbreaks, as critical time is lost during the investigation. Nonetheless, these data streams can be useful in certain scenarios. For instance, Google Flu Trends analyzed massive numbers of Internet search engine queries for syndromic information related to influenza-like illnesses (ILI) [10]. Although the system initially provided promising results that tracked with and even preceded national influenza incidence reporting, changes in the underlying data eventually led to spurious results, ultimately leading to its discontinuation [11].

EHR-based biosurveillance systems benefit from access to substantially more detailed clinical data [12]. Within biosurveillance systems, EHR data is most commonly utilized for *syndromic surveillance*, or the ability to identify non-specific illness clusters before diagnoses are confirmed. Such systems utilize keywords or pattern-matching algorithms derived from selected EHR fields to identify population-level trends or anomalies [13, 14]. Notable US Government-funded operational systems that include syndromic analysis capabilities include the Centers for Disease Control and Prevention’s (CDC) National Syndromic Surveillance Program’s BioSense [15] and the Department of Defense’s (DOD) Electronic Surveillance System for the Early Notification of Community-Based Epidemics (ESSENCE) systems [16, 17]. Although these systems can include anomaly detection capabilities based on case reporting or an aggregate of syndromic information (e.g., clusters of patients with ILI), they do not yet can leverage the complete EHR to inform disease-specific outbreak detection. EHR data has also been used with whole genome sequencing to identify disease transmission routes in hospitals [18, 19] and with structured data and clinical notes to improve case detection from EHR for surveillance [20]. Multiple publications have employed EHR for case detection of confirmed and suspected cases of influenza and COVID-19 for outbreak detection and prognostics [21-24].

The richness of information in modern EHR systems could provide an opportunity to predict the final diagnosis of a patient even before a final diagnosis is recorded. Early symptom data coupled with prescribed medications, orders for laboratory or diagnostic tests, and other clinical data in the EHR can potentially be used to predict the final diagnosis, since these are direct artifacts of a longitudinal clinical workup process during which additional testing and temporal assessments of a disease trajectory are used to reduce diagnostic uncertainty and improve diagnostic accuracy. By exploiting the complete electronic health record of a patient and leveraging recent advances in machine learning-based predictive analysis, we aimed to reduce the time required to identify health anomalies by predicting patient diagnoses before an official diagnosis is recorded in the EHR. Our hypothesis was that aggregating patient-level predicted diagnoses by utilizing extensive EHR clinical data, would enable earlier outbreak detection compared with use of EHR case reporting alone. To this end, we followed a conceptual architecture previously proposed by our research group [25] and developed a two-step process to assess timeliness of outbreak detection: 1) predict individual patient diagnoses and treat high-confidence predictions as new diagnoses; and 2) combine predicted diagnoses with confirmed diagnoses and assess whether spatiotemporal outbreaks could be detected earlier than with confirmed case diagnoses alone. The integration of a spatiotemporal layer is based on the premise that transmissible infectious diseases likely manifest in geographically clustered patterns, and that aggregating high-confidence predictions within defined spatiotemporal unit enhances the signal-to-noise ratio for outbreak detection. **Figure 1** illustrates the workflow, integrating machine learning-based patient diagnosis prediction into EHR-based outbreak detection.

**Figure 1.**
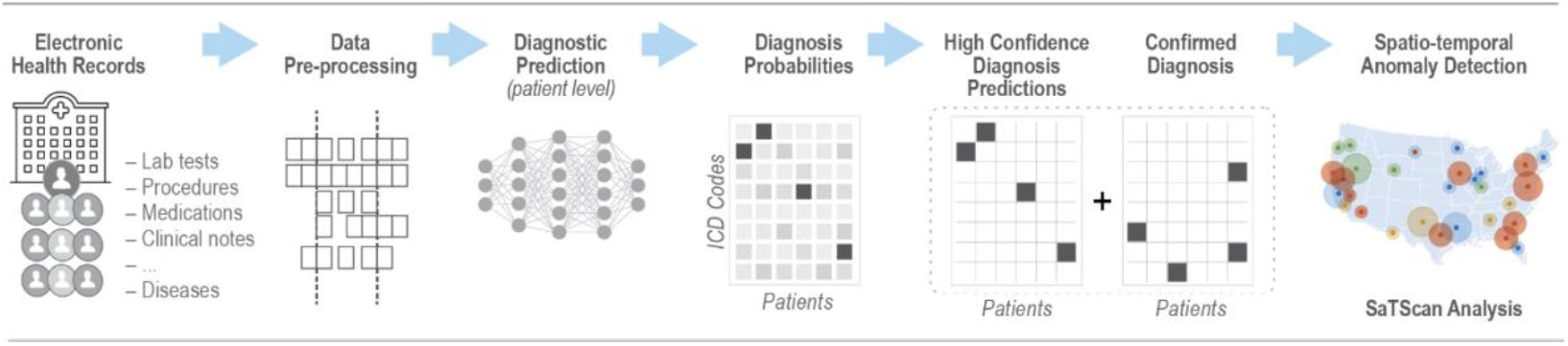
EHR-based machine learning workflow with patient-level diagnosis prediction for infectious disease outbreak detection.

We demonstrate the performance of our proposed framework on a longitudinal EHR dataset of 4.5 million patients from 2010 to 2022 from Kaiser Permanente Northern California, an integrated healthcare delivery system. We considered two types of disease outbreaks, (1) an anomalous spatiotemporal cluster of patients with one of 55 specific infectious disease ICD codes, and (2) an anomalous spatiotemporal cluster of patients with a set of non-specific ICD codes for gastrointestinal (GI), respiratory and neurological diseases. We focused on non-specific diagnoses within the GI, respiratory, and neurologic systems because those were likely to capture the most common conditions presented by patients with a transmissible infectious disease. For example, a patient with a potential meningitis infection may be given a “ not otherwise specified” diagnosis as a placeholder until the point at which a definitive microbiologic etiology is specified. In some instances, a definitive microbiologic pathogen is never identified, and the final diagnosis remains classified with an “ unspecified disease” category.

## Methods

### Data Description

We used EHR data from Kaiser Permanente Northern California (KPNC), which utilizes the Epic EHR software (www.epicsystems.com) across all healthcare settings, including >200 medical clinics and 21 hospitals. The KPNC EHR (KP HealthConnect) employs a common medical record number system for all patient encounters and administrative transactions, including those involving the small fraction of care received outside the system, as well as non-members cared for at KPNC emergency departments and hospitals. Prior work by KPNC describes the integrated information systems and the ability to extract and use clinical, diagnostic, laboratory, and health services data [26-30]. This study was approved by the KPNC Institutional Review Board.

To establish a dataset that provided a year-long sequence of events preceding the relevant diagnosis for each patient, we looked back one year from an index date and generated a data file that combined the following data elements with time stamps, where appropriate: 1) age, sex, race/ethnicity, duration of membership covered over the past one year (a patient could have a gap of up to 3 months and still be considered a member); 2) census tract neighborhood code; 3) vital signs (temperature, systolic BP, diastolic BP, heart rate, respiratory rate); 4) all laboratory test orders; 5) all ICD diagnosis codes; 6) all procedure codes; 7) medication orders (aggregated within pharmaceutical classes); 8) encounter type; 9) emergency department patient chief complaint; and 10) clinical notes turned into a vector representation (embeddings) using the BioClinicalBERT model [31].

The case cohort consists of patients who had infectious disease and unspecified GI, respiratory, and neurological diseases based on ICD diagnosis codes. The ICD codes used for disease categorization are provided in **Tables S1** and **S2** of the supplementary material. The cases consisted of patients within a calendar year who had an ICD diagnosis of interest whose index date was marked by the date that specific diagnosis was assigned. The control cohort consisted of patients from the same population that *did not* have any of the ICD diagnoses in **Tables S1** and **S2**. Eligible controls included all patients not in the targeted disease cohorts within the given matched year. In other words, even if a patient had an infectious disease ICD in 2012, they would be eligible to act as a control in 2011. Additionally, control patients were matched directly on age (+/-5 years), sex, and geocoded location (to facilitate spatiotemporal anomaly detection). Once all possible controls were identified, 2 were randomly selected for each index case. For each of the controls, the same data elements were obtained in the 1-year lookback timeframe. All data were combined and de-identified before being transferred outside of the KPNC system. The summary of the patient demographics and controls for infectious disease and unspecified GI, respiratory, neurological diseases are provided in the supplementary material (**Tables S3, S4, S5**, and **S6**).

### Machine Learning Models for Patient Diagnoses Prediction

Using patients’ historical medical records, demographics, and embedded clinical notes, we trained and compared multiple multi-class classification models for the following diagnostic classification tasks: a) specified infectious diseases and b) unspecified GI/respiratory/neurological diseases. We explored two different model architectures, namely an XGBoost model based on aggregated data and fine-tuned large language Models (LLMs) based on sequential data.

While the class labels for the unspecified disease categories were balanced (see supplementary material **Tables S14, S15** and **Figure S1**), the class labels for infectious diseases were severely imbalanced (see supplementary material **Figure S2**). To mitigate this issue, we applied sample weighting to the loss function across all models implemented in this paper. Each sample was assigned a weight inversely proportional to the frequency of cases of that disease in the training dataset. The weight for the *i*-th sample associated with the *k*-th disease is calculated as:

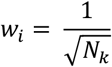

where *N*_*k*_ is the number of cases of the *k-*th infectious disease in the training set [32].

To evaluate the models for diagnostic prediction, while ensuring no leakage between test/train data, we initially trained the models using data from 2010 to 2018 (with 10% of training data being kept aside for model validation and ‘*early stopping*’ to prevent overfitting) and tested on data from 2019. Subsequently, we adopted an ‘*expanding window*’ approach where data from 2019 was incorporated into the training dataset for testing on data from 2020, and so on. This approach allowed the model to be fine-tuned annually to incorporate new data.

### LLM-based predictors

To leverage the sequential nature of EHR data and harness the capabilities of state-of-the-art pre-trained large language models (LLMs), our approach involved fine-tuning several well-known LLMs on structured EHR data. The structured EHR data was pre-processed to align with the expected input format of these LLMs and embeddings were generated for this data. In addition to structured EHR data, we integrated patient demographics and clinical notes in our prediction pipeline by transforming them into embeddings using one-hot-encoding and a pre-trained LLM (BioClinicalBERT model [31]), respectively. All these embeddings (structured sequential data, demographics data and clinical notes) were then aggregated via a late-fusion strategy. This method allowed the model to effectively capture both the structured and unstructured patient data, providing a comprehensive and holistic representation of the patient’s condition. **Figure 2** presents a schematic of our proposed LLM-based disease diagnosis prediction method, illustrating a hypothetical patient diagnosed with “ Monkeypox infection” .

**Figure 2.**
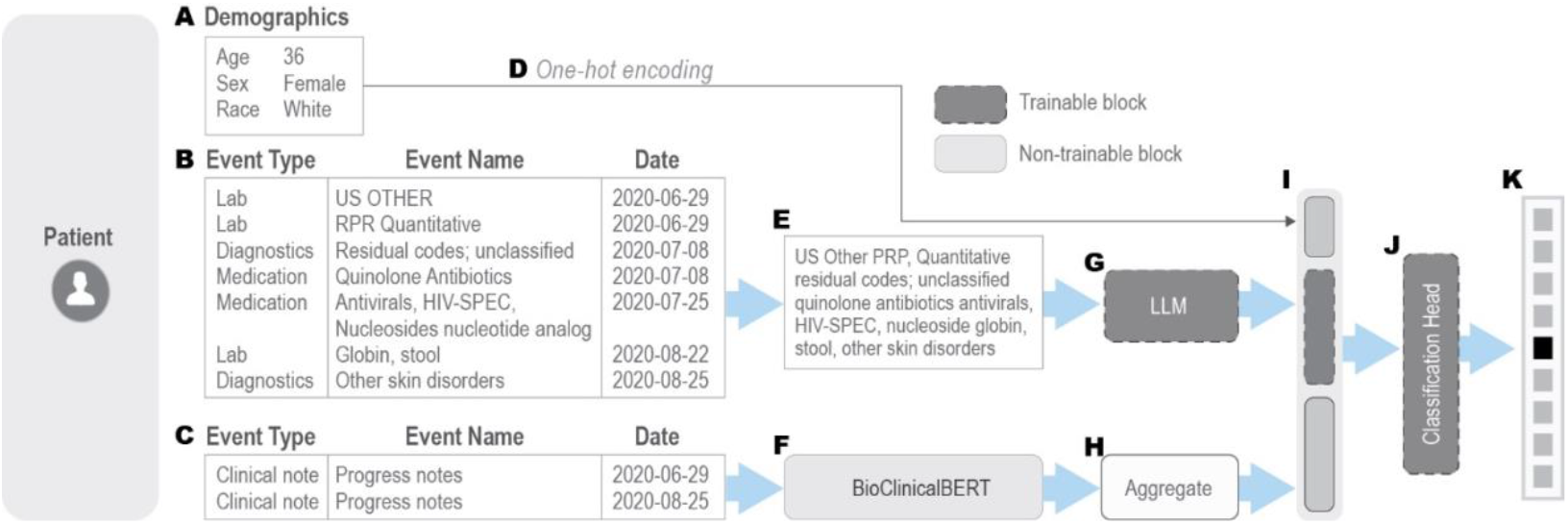
Example of an LLM-based diagnosis prediction with structured EHR data as proposed in this paper. (A) Patient demographics, including age, sex, and race. (B) Medical history within a defined time window (2 months in this case), incorporating medications, lab tests, diagnosis codes, and chief complaints. (C) Clinical notes recorded during the same period. (D) Demographics are one-hot encoded separately and concatenated into a single input vector. (E) Medical events within the window are represented as a concatenated text sequence, omitting event types and dates. (F) Raw clinical notes are processed using a pre-trained BioClinicalBERT model to extract embeddings. (G) The patient’s medical history “ text” is input into a trainable LLM. (H) If multiple clinical notes exist within the window, separate embeddings are extracted and aggregated (e. g., using mean function). If no clinical note is available within the time window, an embedding for the sentence “ No additional clinical information” is provided as input. (I) One-hot encoded demographics, LLM-generated embeddings, and aggregated clinical note embeddings are concatenated in a late fusion fashion. (J) A trainable classification head integrates these inputs. (K) The model outputs probability scores for each disease.

LLMs are pre-trained on large amounts of general text data. However, specialized LLMs are often further trained on domain specific data; for example, LLMs geared for biological tasks are often fine-tuned on biological literature or clinical notes. To understand the impact of pre-training on biological/clinical notes on diagnostic prediction and access if greater alignment between pre-training data and our current task improves transfer learning and, consequently diagnostic prediction performance, we investigated three pre-trained BERT-based LLMs [33], each trained with different amounts of biological/clinical data:

1. **BERT Base Model [33]**, pre-trained on BookCorpus (a dataset of 11,038 unpublished books) and English Wikipedia.
2. **BioBERT [34]**, a fine-tuned version of the BERT Base Model, further trained on PubMed abstracts and full-text articles.
3. **BioClinicalBERT [31]**, a fine-tuned version of BioBERT refined on all notes from MIMIC III [35], a database containing EHR from ICU patients at Beth Israel Hospital in Boston, MA.

These models represent a progression in the volume of biological/clinical data used for training. All three models have approximately 110 million trainable parameters. As shown in **Figure 2**, to encode a patient’s medical history into a format compatible with the pre-trained LLMs, we concatenate the sequence of medical events in a patient’s EHR record (after dropping event type, numerical components such as laboratory tests results, and recorded dates) into a single textual sequence, preserving their chronological order. The model architectures and pre-trained weights for all the three models were obtained through HuggingFace’s Python API (https://huggingface.co).

While newer and bigger GPT-style models are becoming increasingly available, we opted for BERT-based architectures due to their bidirectional encoding, strong performance on discriminative tasks, and extensive benchmarking in the biomedical domain. Additionally, GPT-style models are generally more resource-intensive, making BERT-based models a more practical and interpretable choice for our fine-tuning objectives. In future work, we plan to explore the performance and applicability of other LLM architectures on this task.

During the fine-tuning phase for our specific downstream diagnostic prediction task, the pre-trained LLM’s original masked-token prediction head was replaced with an untrained multiclass softmax head, configured for diagnostic prediction outcomes. The pre-trained layers of the LLM were allowed to update during training. We used a learning rate *lr*=5e-5 and trained the models for a maximum of 20 epochs, employing early stopping based on performance on a validation set.

### Count-based predictor

An alternative approach to modeling sequences involves aggregating sequential information in a pre-determined time window into a fixed-length vector which can then be used as input feature for traditional machine learning models like Random Forest or XGBoost. This approach, explored in prior studies [36, 37], has demonstrated strong predictive performance. A commonly used aggregation function is “ count”, where the fixed-length vector corresponds to the total number of potential medical events (e.g., labs, medications, procedures). Each element in the array represents the frequency of a specific event observed in the patient’s historical medical record. The resulting vector is typically high-dimensional and sparse, reflecting the large dictionary of possible medical events, with only a subset occurring in practice. In our dataset, the structured EHR count vector had a dimensionality of 4,818, covering lab tests (2,565), medications (681), vital sign measurements (7), diagnosis codes (284), and chief complaints (1,281). Similar to the LLM-based prediction models, patient demographics are one-hot encoded and concatenated with clinical note embeddings and the medical event count vector. This combined input is then fed into our chosen ML model, an XGBoost multiclass classifier. The final input vector has 5,599 dimensions, consisting of 4,818 from the EHR count vector, 13 from one-hot encoded demographics, and 768 from clinical note embeddings. The choice of XGBoost is motivated by the robust performance of XGBoost in healthcare related classification problems [38, 39]. As discussed later in the Results section, besides serving as a robust baseline for the LLM based approaches, XGBoost is particularly effective for detecting certain infectious diseases. We hypothesize that these infectious diseases may be associated with very specialized labs or medications, which a decision-tree based model such as XGBoost, is able to recognize easily.

### Evaluation Metrics

To evaluate the performance of the machine learning models for diagnostic prediction, we considered several binary and multi-class classification metrics including Micro and Macro F1-Scores, Macro Balanced Accuracy and Matthew Correlation Coefficient (MCC). These metrics look at different aspects of the model performance. Additional details of the evaluation metrics are provided in the supplementary material (**Table S7**). In our analysis, we used the scikit-learn (https://scikit-learn.org/stable/) Python package implementation of these metrics.

### Spatiotemporal outbreak detection

Spatiotemporal scan statistics is a commonly used methodology for investigating spatiotemporal anomalies in disease outbreak data [40-43]. With scan statistics, an anomaly score is computed over different spatiotemporal windows, and the window with the highest statistically significant score is flagged as the time/location of an outbreak. Briefly, anomaly detection is framed as a hypothesis testing problem. Under the null hypothesis H0, there is no anomaly, and the incidence of the target disease comes from the same probability distribution regardless of space and time. Under the alternative hypothesis H1(S, T), there is a spatial cluster S and temporal window T where the target disease is generated from a probability distribution with higher mean than in all other windows. The goal in spatiotemporal scan statistics is to find the cluster (S, T) that maximizes the likelihood ratio—i.e., test statistic—F(S, T) given the reported case counts C:

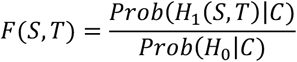

The statistical significance of the highest scoring cluster is assessed by computing the recurrence interval, defined as the inverse of the probability of observing a spatiotemporal cluster with a test statistic at least as extreme as the one detected, under the null hypothesis of no clustering. This reflects the expected time between such events occurring by chance and is typically estimated via Monte Carlo simulation. In this study, a recurrence interval threshold of 100 years was used to define statistically significant spatiotemporal clusters, corresponding to events expected to occur by chance no more than once per century under the null hypothesis.

We simulated a prospective study by performing spatiotemporal scan statistics analysis in 24-hour increments from January 1, 2014 to December 31, 2022 using the SaTScan™ software (https://www.satscan.org/) for spatiotemporal outbreak detection. A configuration file listing all the parameters for our analysis can be found as part of the supplementary material (**Table S17**).

To determine the existence of a disease outbreak on a given day *x*, we considered the total reported case counts (based on diagnosis codes recorded in EHR) up to day *x* in each census tract. Moreover, we made diagnosis predictions using the models described above on all EHR data available before day *x*. We trained ML models with data from 2010 up to the end of each year from 2013 to 2022. When making predictions for a day in a year *y*, we used the model trained up to year (*y*-1). The number of predictions with score greater than a disease-specific threshold was added to the reported counts on day *x*. We refer to this number as the *predicted* case count.

The primary metric for evaluating the effectiveness of diagnostic predictions in enhancing early disease outbreak detection was the outbreak lead time. Lead time was defined as the interval, in days, between the detection of an outbreak utilizing both predicted and reported case counts, and its detection relying solely on reported case counts. This analysis was conducted at a spatial resolution of census tracts and a temporal resolution of one day. Consequently, detected outbreaks manifested as spatiotemporal clusters encompassing one or more contiguous census tracts within Northern California over a duration of one or more full days. Consistent with public health priorities, the study focused on geographically and temporally compact outbreaks; therefore, the analysis was constrained to identify clusters comprising no more than 50% of the available census tracts and spanning a maximum duration of 28 days.

The performance characteristics of our outbreak detection algorithm, particularly false positive rates, and outbreak detection lead times, are highly sensitive to the disease-specific thresholds employed. Conventional detectors aim to control an error rate, such as the False Discovery Rate (FDR) (defined as the ratio of false positives to the total number of detections) below a predefined maximum while optimizing the true positive rate (statistical power). However, the low frequency of documented outbreaks for individual diseases during the 2010-2022 study period (ranging from 0 to 20 occurrences) precludes the robust estimation of FDR using standard methodologies. To ensure minimum data quality, first the evaluation of detection lead times was restricted to diseases with a minimum of five confirmed outbreaks within the study timeframe. Subsequently, this study implemented a threshold calibration strategy of controlling the False Discovery Rate (FDR) at or below 1/6, which corresponds to a maximum of one false positive outbreak occurrence for a disease with a total of 5 true outbreaks over the entire study period of 9 years. Prospective real-world deployment would necessitate threshold recalibration based on more extensive data to facilitate reliable error rate estimation.

Furthermore, the accurate classification of potential false positives presented challenges. A conservative approach was adopted, classifying detections as false positives even in ambiguous situations. For instance, certain scenarios arose where detection statistics incorporating predicted counts marginally surpassed the outbreak classification threshold. Although the corresponding statistics based solely on confirmed cases failed to meet this threshold, underlying positive trends in reported cases suggested a potential nascent outbreak that lacked sufficient reported evidence for confirmation under the established criteria.

For conciseness, the main results report only the maximum lead time achieved by either the XGBoost or BioClinicalBERT model for each condition. Detailed model-specific results are provided in the supplementary material (**Table S16**). In summary, XGBoost predictions generally yielded superior lead times for specific infectious diseases, whereas BioClinicalBERT demonstrated stronger performance for predicting trends in unspecified disease categories.

## Results

We trained several machine learning models for patient-level diagnosis prediction, including BERT, BioBERT, BioClinicalBERT, and XGBoost. The link to each pre-trained LLM used in this work is presented in **Table S8** in the supplementary material. Each model takes as input all clinical events from the two months preceding the disease diagnosis. Although longer observation periods were also evaluated, no significant improvement in classification performance was noted, likely due to the relatively short incubation period of most diseases considered in this paper.

Control patients were included in the training, with each diseased patient matched to two control patients using a two-month observation window aligned with the diseased patient’s window. Controls without any interaction with the healthcare system in that period were excluded. A “ control” class was added alongside the disease classes, resulting in a 56-label multiclass classification problem for infectious diseases and a 5-label multiclass classification problem for unspecified diseases (“ control”, “ known disease”, “ unspecified GI”, “ unspecified respiratory”, and “ unspecified neuro”).

### Infectious disease classification problem

Table 1 shows the mean performance metrics across evaluation years 2019-2022, with separate models trained for each year using cumulative historical data. Results for every year available in the dataset are presented in **Table S9** in the supplementary material. Analysis includes infectious diseases with 5 or more confirmed cases in the test year. Diseases with fewer than 5 cases were excluded, as the sample size is insufficient for training and evaluation. After that, the total number of infectious diseases reduced from 55 to 32, plus the additional class for “ control” . The list of infectious diseases is provided in the supplementary material (**Table S1**).

Rare diseases present a unique challenge for predictive modeling due to their inherently low prevalence, which limits the amount of data available for training and evaluation. However, these diseases are of critical importance for biosurveillance, as their outbreaks, though infrequent, can pose significant risks to public health and may require swift and effective response strategies. To assess the robustness of each model in handling rare diseases, we computed the same performance metrics as for the broader dataset but focused specifically on the 15 rarest diseases in our data. The list of 15 rarest diseases is provided in the supplementary material (**Table S1**). **Table 2** provides detailed comparison of the models’ performance, highlighting their ability to detect and predict these challenging cases. This analysis emphasizes the importance of developing models capable of addressing the unique demands of rare disease surveillance.

**Table 1.** Average and standard deviation classification performance of machine learning models for patient-level infectious disease diagnosis prediction, evaluated over the years 2019 to 2022, using regular unweighted (Unwtd) and sample weighted (Wtd) loss functions. It includes all diseases that have at least 5 confirmed cases in the test year. All metrics range from 0 to 1, with higher values indicating better performance and an ideal score of 1.

|  | F1-Micro |  | F1-Macro |  | MCC |  | BA Macro |  |
| --- | --- | --- | --- | --- | --- | --- | --- | --- |
|  | Unwtd | Wtd | Unwtd | Wtd | Unwtd | Wtd | Unwtd | Wtd |
| **XGBoost** | 0.81<br>(0.012) | **0.78**<br>**(0.015)** | 0.40<br>(0.03) | **0.38**<br>**(0.02)** | 0.63<br>(0.03) | 0.61<br>(0.03) | 0.64<br>(0.016) | **0.69**<br>**(0.017)** |
| **BERT** | 0.81<br>(0.012) | 0.76<br>(0.013) | **0.44**<br>**(0.04)** | 0.31<br>(0.01) | 0.65<br>(0.02) | 0.59<br>(0.02) | 0.63<br>(0.013) | 0.66<br>(0.005) |
| **BioBERT** | 0.81<br>(0.011) | 0.76<br>(0.008) | 0.42<br>(0.03) | 0.29<br>(0.02) | 0.64<br>(0.02) | 0.58<br>(0.02) | 0.63<br>(0.027) | 0.67<br>(0.009) |
| **BioClinicalBERT** | **0.82**<br>**(0.008)** | **0.78**<br>**(0.018)** | 0.42<br>(0.02) | 0.34<br>(0.02) | **0.67**<br>**(0.02)** | **0.62**<br>**(0.03)** | **0.65**<br>**(0.008)** | **0.69**<br>**(0.01)** |

**Table 2.** Average and standard deviation classification performance of machine learning models for patient-level infectious disease diagnosis prediction, evaluated over the years 2019 to 2022, using regular unweighted (Unwtd) and sample weighted (Wtd) loss functions. Analysis focuses on the 15 least common infectious diseases, each with at least 5 confirmed cases in the test year. Performance metrics are scaled from 0 to 1, where 1 represents perfect prediction accuracy.

|  | F1-Micro |  | F1-Macro |  | MCC |  | BA Macro |  |
| --- | --- | --- | --- | --- | --- | --- | --- | --- |
|  | Unwtd | Wtd | Unwtd | Wtd | Unwtd | Wtd | Unwtd | Wtd |
| **XGBoost** | **0.18**<br>**(0.05)** | 0.29<br>(0.04) | 0.35<br>(0.05) | **0.45**<br>**(0.03)** | **0.20**<br>**(0.05)** | 0.29<br>(0.03) | **0.59**<br>**(0.03)** | **0.65**<br>**(0.02)** |
| **BERT** | 0.14<br>(0.05) | 0.24<br>(0.04) | **0.45**<br>**(0.07)** | 0.39<br>(0.06) | 0.14<br>(0.05) | 0.24<br>(0.04) | 0.57<br>(0.02) | 0.62<br>(0.013) |
| **BioBERT** | 0.12<br>(0.08) | 0.27<br>(0.08) | 0.40<br>(0.06) | 0.39<br>(0.02) | 0.12<br>(0.08) | 0.26<br>(0.03) | 0.56<br>(0.04) | 0.63<br>(0.007) |
| **BioClinicalBERT** | 0.18<br>(0.04) | **0.31**<br>**(0.04)** | 0.44<br>(0.05) | **0.45**<br>**(0.04)** | 0.18<br>(0.04) | **0.30**<br>**(0.05)** | 0.58<br>(0.01) | **0.65**<br>**(0.03)** |

Table 3 shows model performance in terms of balanced accuracy for each individual disease (and control) for both unweighted and weighted strategies. A similar table for the F1-score performance metric is the supplementary material (**Table S10**).

**Figure** shows model performance (F1-score) vs. the number of cases available for training for each infectious disease for the BioClinicalBERT model.

**Table 3.** Average Balanced Accuracy per disease over the years of 2019 through 2022, comparing Unweighted and Weighted loss strategies. The number of cases for each disease, spanning from 2010 to 2022, is shown next to the disease name. Diseases are sorted by total case count. The best performing model (closest to 1) for each strategy and disease is highlighted in pink. Diseases for which all models performed no better than random (Balanced Accuracy = 0. 5) are highlighted in yellow. * There are no cases of Monkeypox prior to 2022 and all 488 cases for Monkey Pox occurred in 2022.

| Disease (No. cases) | Unweighted |  |  |  | Weighted |  |  |  |
| --- | --- | --- | --- | --- | --- | --- | --- | --- |
|  | XGBoost | BERT | BioBERT | Bio-ClinicalBERT | XGBoost | BERT | BioBERT | Bio-ClinicalBERT |
| Control (131,202) | 0.84 | 0.89 | 0.90 | 0.90 | 0.86 | 0.89 | 0.88 | 0.90 |
| Syphilis (17,748) | 0.83 | 0.86 | 0.86 | 0.86 | 0.85 | 0.84 | 0.84 | 0.85 |
| Campylobacteriosis (8,445) | 0.80 | 0.83 | 0.80 | 0.82 | 0.78 | 0.79 | 0.76 | 0.79 |
| Chickenpox (6,671) | 0.73 | 0.79 | 0.79 | 0.79 | 0.78 | 0.79 | 0.79 | 0.79 |
| Pertussis (6,131) | 0.81 | 0.87 | 0.87 | 0.86 | 0.87 | 0.89 | 0.87 | 0.89 |
| Tuberculosis (6,060) | 0.73 | 0.71 | 0.72 | 0.74 | 0.76 | 0.69 | 0.70 | 0.72 |
| Salmonellosis (5,279) | 0.70 | 0.69 | 0.70 | 0.73 | 0.71 | 0.69 | 0.67 | 0.71 |
| Shigellosis (3,474) | 0.59 | 0.58 | 0.58 | 0.54 | 0.64 | 0.61 | 0.64 | 0.62 |
| Meningitis (2,872) | 0.82 | 0.77 | 0.75 | 0.81 | 0.85 | 0.75 | 0.73 | 0.77 |
| Haemophilus (1,624) | 0.67 | 0.70 | 0.68 | 0.75 | 0.75 | 0.75 | 0.74 | 0.78 |
| Hepatitis A (1,239) | 0.53 | 0.53 | 0.53 | 0.56 | 0.59 | 0.60 | 0.57 | 0.58 |
| Monkeypox* (488) | 0.50 | 0.50 | 0.50 | 0.50 | 0.50 | 0.50 | 0.50 | 0.50 |
| Dengue Virus Infection (480) | 0.56 | 0.61 | 0.57 | 0.62 | 0.57 | 0.61 | 0.62 | 0.65 |
| Hemolytic Uremic Syn. (385) | 0.56 | 0.51 | 0.53 | 0.53 | 0.61 | 0.60 | 0.62 | 0.58 |
| Legionellosis (363) | 0.58 | 0.50 | 0.52 | 0.57 | 0.59 | 0.54 | 0.57 | 0.61 |
| Malaria (354) | 0.63 | 0.66 | 0.64 | 0.66 | 0.71 | 0.66 | 0.67 | 0.69 |
| West Nile Virus (324) | 0.53 | 0.51 | 0.52 | 0.53 | 0.57 | 0.53 | 0.56 | 0.56 |
| E. coli (317) | 0.50 | 0.50 | 0.50 | 0.50 | 0.52 | 0.54 | 0.52 | 0.57 |
| Typhoid Fever (282) | 0.52 | 0.50 | 0.51 | 0.52 | 0.59 | 0.54 | 0.57 | 0.56 |
| Relapsing Fever (278) | 0.55 | 0.53 | 0.53 | 0.55 | 0.61 | 0.60 | 0.58 | 0.59 |
| Cryptosporidiosis (248) | 0.75 | 0.77 | 0.68 | 0.77 | 0.79 | 0.79 | 0.77 | 0.79 |
| Q Fever (221) | 0.79 | 0.74 | 0.74 | 0.83 | 0.83 | 0.74 | 0.76 | 0.81 |
| Poliovirus Infection (195) | 0.58 | 0.50 | 0.50 | 0.50 | 0.65 | 0.51 | 0.52 | 0.59 |
| Rubella Synd. (174) | 0.51 | 0.50 | 0.50 | 0.50 | 0.65 | 0.55 | 0.63 | 0.65 |
| Trichinosis (172) | 0.57 | 0.69 | 0.68 | 0.72 | 0.71 | 0.80 | 0.83 | 0.83 |
| Mumps (167) | 0.52 | 0.53 | 0.54 | 0.57 | 0.53 | 0.62 | 0.63 | 0.67 |
| Tetanus (142) | 0.62 | 0.57 | 0.50 | 0.61 | 0.63 | 0.64 | 0.59 | 0.67 |
| Listeriosis (135) | 0.50 | 0.50 | 0.50 | 0.50 | 0.53 | 0.54 | 0.56 | 0.56 |
| Meningococcal Infect. (126) | 0.50 | 0.50 | 0.50 | 0.50 | 0.50 | 0.50 | 0.50 | 0.50 |
| Creutzfeldt-Jacob Dis. (106) | 0.58 | 0.50 | 0.50 | 0.50 | 0.69 | 0.50 | 0.54 | 0.59 |
| Measles (Rubeola) (103) | 0.50 | 0.50 | 0.50 | 0.50 | 0.50 | 0.50 | 0.50 | 0.50 |
| Rickettsial Diseases (99) | 0.50 | 0.50 | 0.50 | 0.50 | 0.56 | 0.50 | 0.56 | 0.50 |
| Chikungunya Virus Infec. (97) | 0.50 | 0.50 | 0.50 | 0.50 | 0.50 | 0.56 | 0.50 | 0.50 |
| Babesiosis (92) | 0.50 | 0.50 | 0.50 | 0.50 | 0.50 | 0.61 | 0.60 | 0.58 |
| Brucellosis (76) | 0.51 | 0.50 | 0.50 | 0.50 | 0.50 | 0.56 | 0.56 | 0.56 |
| Zika Virus Infection (75) | 0.53 | 0.50 | 0.50 | 0.50 | 0.53 | 0.70 | 0.65 | 0.71 |
| Botulism (65) | 0.64 | 0.50 | 0.50 | 0.50 | 0.69 | 0.60 | 0.62 | 0.63 |
| Vibrio Infections (65) | 0.50 | 0.50 | 0.50 | 0.50 | 0.50 | 0.58 | 0.58 | 0.58 |
| Rabies (56) | 0.92 | 0.50 | 0.50 | 0.50 | 0.50 | 0.58 | 0.91 | 0.92 |
| Scombroid Fish Poison. (43) | 0.61 | 0.50 | 0.50 | 0.50 | 0.75 | 0.75 | 0.79 | 0.75 |
| Ehrlichiosis (30) | 0.50 | 0.50 | 0.50 | 0.50 | 0.50 | 0.50 | 0.50 | 0.50 |

To further characterize our modeling results we conducted an ablation study to evaluate the discriminative power of each medical data type: vital signs, medication, off-target diagnoses, and lab tests. Chief complaint data was excluded from the ablation study, as it is limited and available for only a small subset of subjects. Detailed tables are provided in the supplementary material (**Tables S11** and **S12**). The overall findings indicate that, across all performance metrics, medication is the most informative modality, followed by lab tests and off-target diagnosis codes. Vital signs were the least informative but contributed to improved overall performance when combined with all other data types, which is unsurprising since vital signs are non-specific with abnormalities shared across many diseases and conditions.

### Unspecified disease classification problem

In addition to the infectious disease classification problem, we examined the “ unknown” or “ not otherwise specified” set of ICD codes and defined secondary disease labels such as “ unspecified GI”, “ unspecified respiratory”, and “ unspecified neurological” diseases. The details regarding the ICD codes associated with the unspecified diseases are provided in the supplementary material (**Table S2**). The average diagnostic performance results over the years of 2019 to 2022 for unspecified diseases for XGBoost and fine-tuned BioClinicalBERT are provided in **Table 4**. Results for all evaluation years (2014 through 2022) considering all four classification performance metrics are available in **Table S13. Tables S14** and **S15** provide details on the number of training and evaluation samples for each unspecified disease for every training period, as well as for the “ control” and “ known disease” groups.

**Table 4.** Average and standard deviation of multiclass classification performance of machine learning models for unspecified disease classification prediction, evaluated over the years 2019 to 2022, using XGBoost and BioClinicalBERT models. Both models were trained with sample-weighted loss function (inverse square root of class frequency). Performance metrics are scaled from 0 to 1, where 1 represents perfect prediction accuracy.

| Model | F1-Micro | F1-Macro | MCC | BA Macro |
| --- | --- | --- | --- | --- |
| **XGBoost** | 0.72 (0.002) | 0.51 (0.01) | 0.45 (0.01) | 0.71 (0.01) |
| **BioClinicalBERT** | **0.75 (0.006)** | **0.57 (0.01)** | **0.53 (0.01)** | **0.75 (0.01)** |

### Spatiotemporal outbreak detection

In **Table 6**, we report the number of outbreaks, average lead time for events with positive lead time for infectious diseases with at least five outbreaks, as well as the unspecified GI and respiratory diseases. For the infectious diseases, the lead time ranges from 2.5 days, in the case of Shigellosis to over 3 weeks for Syphilis. Notably, lead time aligns with the incubation period of the disease. Syphilis has an average incubation period of 21 days, whereas faster-acting bacterial infections like salmonellosis and shigellosis may develop over the course of hours to days. There were two diseases, rubella syndrome and mumps, with detectable outbreaks that did not yield positive lead times. These diseases have relatively small sample sizes (324 and 174, respectively), which makes it challenging to train accurate predictive models. The overall system performance over the 9-year study period resulted in 12 false positives or 1.33 false positives per year.

**Table 5.** Average and standard deviation Balanced Accuracy score of XGBoost and BioClinicalBERT models per disease over the years of 2019 to 2022) Performance metrics are scaled from 0 to 1, where 1 represents perfect prediction accuracy.

| Model | Control | Known | Unk-GI | Unk-Neuro | Unk-Resp |
| --- | --- | --- | --- | --- | --- |
| **XGBoost** | 0.77 (0.006) | 0.62 (0.009) | 0.70 (0.013) | 0.62 (0.015) | 0.85 (0.019) |
| **BioClinicalBERT** | **0.81 (0.009)** | **0.67 (0.013)** | **0.72 (0.019)** | **0.66 (0.01)** | **0.88 (0.01)** |

**Table 6.** Average lead time and maximum allowable false discovery rate for spatiotemporal outbreak detection using predicted case diagnoses combined with confirmed case diagnoses, as compared to outbreak detection using confirmed case diagnoses alone. *The threshold for calculating the number of false detections was intentionally set to at least 1 during our study period.

| Infectious Disease<br>(No. of diagnosed cases from 2010 to 2022) | Number of true outbreaks/Number of outbreaks detected with a positive lead time | Average positive lead time (days) | Number of false detections* | Average incubation period |
| --- | --- | --- | --- | --- |
| Syphilis (17,748) | 17/20 | 23.8 | 4 | 21 |
| Campylobacteriosis (8,445) | 2/8 | 12.0 | 1 | 4 |
| Pertussis (6,131) | 3/8 | 7.3 | 1 | 8 |
| Tuberculosis (6,060) | 5/7 | 11.2 | 1 | 49 |
| Salmonellosis (5,279) | 2/7 | 5.5 | 1 | 2 |
| Shigellosis (3,474) | 2/8 | 2.5 | 1 | 3 |
| West Nile Virus (324) | 2/5 | 9.5 | 1 | 8 |
| Rubella Synd. (174) | 0/7 | — | 0 | 15 |
| Mumps (167) | 0/5 | — | 0 | 17 |
| **Unspecified Diseases** |  |  |  |  |
| GI (41,146) | 2/11 | 1.0 | 1 | Unspecified |
| Respiratory (25,557) | 2/13 | 1.5 | 1 | Unspecified |

Unspecified diseases have shorter lead times, averaging 1 day for GI diseases and 1.5 days for respiratory diseases. This is likely due to the generic nature of the ICD codes used to classify unspecified diseases, such as “ unknown” or “ not otherwise specified.” These codes are typically assigned at the time of initial symptom presentation, leaving less opportunity for early prediction. Unspecified neurological diseases were excluded from the table due to the absence of outbreaks during the study period.

We chose to only report the lead times when the value was positive. If the system described here were used in an operational context, public health professionals would have knowledge of both whether a spatiotemporal outbreak could be detected via case counts alone, or if the outbreak is instead predicted based on diagnostic prediction information. When there is no lead time, no benefit is gained from use of diagnostic predictions; however, for scenarios where an outbreak is predicted, the average positive lead time is a useful statistic. In other words, the public health official will know the expected outbreak lead time, on average, for that specific disease and can act accordingly.

## Discussion

This study developed machine learning models, trained using extensive electronic health record (EHR) data, to evaluate their potential for accelerating the detection of spatiotemporal infectious disease outbreaks. In our simulated prospective analysis, we observed that incorporating these predictive models yielded outbreak detection lead times up to 23.8 days earlier compared to baseline methods (**Table 6**). Prediction accuracy demonstrated a correlation with the number of confirmed cases available for model training (**Figure 3**). Accordingly, we anticipate that as the availability of EHR data increases, particularly for rare diseases, model performance will improve, potentially leading to enhanced predictive accuracy and longer detection lead times across a broader spectrum of diseases. Such models represent a potentially valuable tool for public health biosurveillance programs, offering the significant advantage of utilizing existing data resources without necessitating additional data collection, and could allow them to balance and combine these insights with those gained from traditional biosurveillance tools.

**Figure 3:**
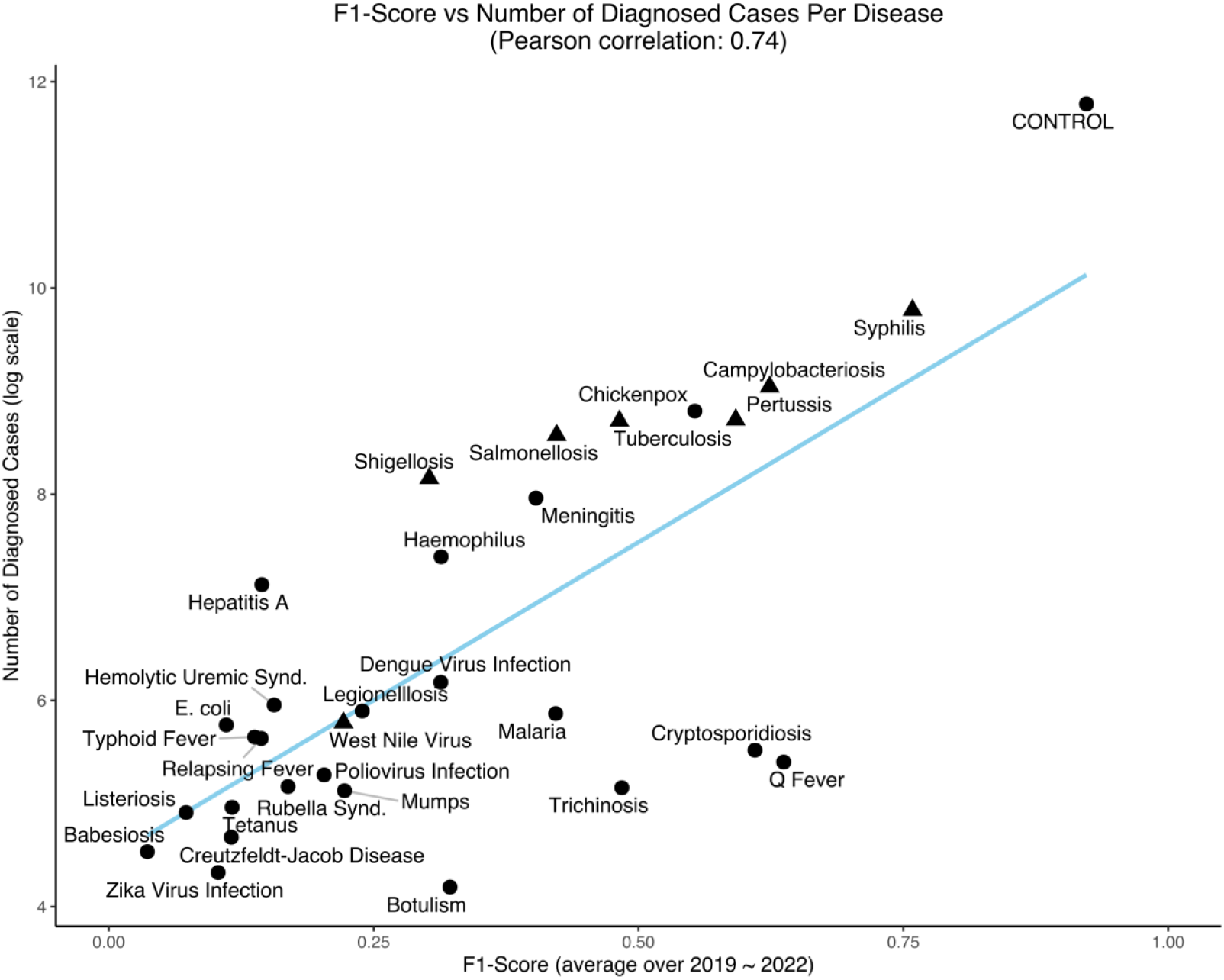
Average F1-score (2019–2022) vs. the number of diagnosed cases (available training samples) in log scale for all infectious diseases with at least 10 cases over the period of 2019-2022 for the BioClinicalBERT model. (▴) indicates (n=7) diseases with a positive lead time for early outbreak detection corresponding with results from Table 6.

Our final model demonstrated the capacity for early outbreak detection across a spectrum of infectious diseases. Notably, future operational implementation of such a model holds significant potential relevance for public health surveillance of GI disease outbreaks. The model enabled the detection of outbreaks attributed to *Campylobacter* spp., *Salmonella* spp., *Shigella* spp., and even unspecified GI illnesses, with lead times ranging from 1 to 12 days compared to conventional detection methods. This early detection capability may prove particularly significant given the typically short incubation periods (2 to 4 days) characteristic of these diseases. Such a detection advantage could provide a crucial window for public health officials to initiate investigations and deploy potentially life-saving interventions, if a common source of the pathogen can be rapidly identified.

Overall, all evaluated models demonstrated predictive capability in this complex multi-class classification task, despite the limited number of training samples available for many diseases (**Table 1**). While the well-known XGBoost with count-based features achieved slightly lower yet comparable performance relative to the language models, the BioClinicalBERT LLM exhibited superior performance across three of the four evaluation metrics employed. This enhanced performance can likely be attributed to BioClinicalBERT’s extensive pre-training on clinical corpora, suggesting that domain-specific transfer learning confers an advantage in predictive capability. The observed substantial disparity between F1-Micro and F1-Macro scores reflects the inherent class imbalance characteristic of disease prevalence data. The F1-Micro metric, being weighted by class sample size, assigns greater importance to performance on more prevalent diseases, whereas the F1-Macro metric accords equal weight to all classes irrespective of their prevalence suggesting that domain-specific transfer learning confers an advantage in predictive capability. Employing sample-weighted loss functions, where weights are proportional to class sizes during model training, revealed a notable trade-off in performance outcomes. Metrics susceptible to class imbalance, namely F1-Micro and Matthews Correlation Coefficient (MCC), exhibited a marked decrease as the training process increasingly prioritized minority classes. These classes typically present greater classification challenges owing to their limited representation in the training data. Conversely, metrics less sensitive to class imbalance, such as F1-Macro and Macro-averaged Balanced Accuracy (BA-Macro), generally demonstrated improvement under this weighted training regimen. This divergence in metric performance became particularly pronounced during the analysis of rare diseases. Specifically, when weighted training was applied, a significant enhancement in classification performance for rare diseases was observed across nearly all models and metrics (**Table 2**). This improvement was particularly evident for metrics more sensitive to class imbalance (F1-Micro and MCC). These findings suggest that implementing weighted training strategies is advantageous when the primary objective is optimizing performance on rare diseases, although this may entail a modest reduction in classification accuracy for more common conditions.

There is no single best-performing model for all or even a majority of diseases. Each group of models, tree-based and LLM-based, has its strengths, suggesting that a combination or ensemble of models is likely the most effective solution for an outbreak detection system (**Table 3**). For certain diseases where a single lab test or medication determines whether a subject has the disease, tree-based models are likely to perform better, as these models excel at identifying and prioritizing the most predictive features. In contrast, for diseases where the diagnostic process depends on the sequence of events, LLM-based models are better equipped to extract discriminative information. Importantly, some conditions (e.g., diarrheal diseases) may trigger the same diagnostic workup which may hamper precise prediction of a specific pathogen but still highlight anomalies in the typical occurrence of those conditions within spatiotemporal units and facilitate investigation. Aside from a few exceptions, we observe that the models perform better for diseases with a larger number of cases, and consequently more training samples, while performance declines for rarer diseases. However, it is noticeable that model performance significantly improves on rarer diseases when sample weighted training strategy is used, at the cost of a small decrease in performance for diseases with larger number of cases. For some diseases such as Listeriosis, Rickettsial Diseases, Chikungunya Virus Infection, Babesiosis and Vibrio Infections, where none of the models produced meaningful predictions under the unweighted strategy, we observe the emergence of predictive power under the weighted strategy.

For unspecified disease classification, **Table 4** highlights that in general, a fine-tuned BioClinicalBERT significantly outperforms an XGBoost model on all metrics. Unlike infectious disease classification, where 50% of the diseases have fewer than 130 samples (25th percentile: 34, 50th percentile: 126, 75th percentile: 359) for over the entire period considered, from 2010 to 2022, unspecified diseases on the other hand have significantly larger sample sizes— 22,695 for the least frequent class, unspecified neuro. The increased availability of training data, as provided in **Tables S14** and **S15**, particularly benefits flexible models like LLMs, potentially explaining why fine-tuned BioClinicalBERT outperforms XGBoost for this classification task. Given their greater complexity, fine-tuned LLMs benefit from larger sample sizes, which enhance both training and generalization. There is a large disparity in model performance across various unspecified diseases (**Table 5**). Both models perform well for unspecified respiratory disease, which may be due to the presence of specific markers that are commonly associated with the workup of respiratory conditions prior to definitive diagnosis. On the other hand, the performance of both models for unspecified neurological disease is poor, which may be attributed to both the less common occurrence of unspecified neurologic disease as well as the significantly more complex diagnostic process for neurologic disease. The subpar performance of both models for the “ known disease” class may be attributed to the fact that this class is assigned a very wide range of ICD codes (all ICD codes except those in **Table S2**) and is very heterogeneous and complex.

A correlation was observed between the lead time for outbreak detection and the duration of the corresponding disease incubation period. However, the sample size was insufficient for robust statistical analysis. This relationship is theoretically anticipated, as the potential for earlier diagnosis in diseases with longer incubation periods provides a wider window for diagnosis, which consequently may extend the outbreak detection lead time at the population level. Furthermore, we found that ML model performance exhibited a strong positive correlation (Pearson correlation coefficient = 0.74) with the volume of available training data (**Figure 3**). While this finding aligns with established principles of machine learning, it highlights the potential for significant model improvement through training on EHR data from larger cohorts. This is particularly pertinent for rare diseases, where the models demonstrated limited diagnostic accuracy. This limitation is likely attributable not to inherent diagnostic challenges, but rather to inadequate training data, which restricts the ML algorithms’ ability to learn the unique features associated with these conditions and their clinical diagnostic pathways.

Future research conducted by our team will investigate data augmentation approaches and other methodologies to enhance diagnostic accuracy for rare diseases. For instance, the impact on system performance of combining EHR data collected from one health organization with data from a second organization to augment the available training data for diagnostic prediction of rare diseases is not yet understood. Similarly, methodological approaches, such as the integration of biological knowledge graphs in our prediction pipeline warrant further evaluation to improve system performance for rare diseases.

## Disclaimer

This work was performed under the auspices of the US Department of Energy by Lawrence Livermore National Laboratory (LLNL) under contract DE-AC52-07NA27344. LLNL-JRNL-2005606. The views and conclusions contained in this document are those of the authors and should not be interpreted as necessarily representing the official policies, either expressed or implied, of DHS or the U.S. Government. The DHS does not endorse any products or commercial services mentioned in this presentation. In no event shall the DHS have any responsibility or liability for any use, misuse, inability to use, or reliance upon the information contained herein. In addition, no warranty of fitness for a particular purpose, merchantability, accuracy, or adequacy is provided regarding the contents of this document.

## Data Availability

The datasets generated during and/or analyzed in this report are the property of Kaiser Foundation Health Plan, Inc. and are not publicly available due to their potentially identifiable information and Kaiser Permanente Northern California privacy regulations.

## Code Availability

The code is available to editors and reviewers upon request and will be publicly released to the community following the paper’s acceptance.

