## Supplementary material for "AI-Enabled Diagnostic Prediction within Electronic Health Records to Enhance Biosurveillance and Early Outbreak Detection": SI

**Table of Contents**

Table S1. ICD codes for infectious disease categorization

Table S2. ICD codes for unspecified disease categorization

Table S3. Demographic characteristics of the infectious disease cohort

Table S4. Demographic characteristics of the unspecified gastrointestinal disease cohort

Table S5. Demographic characteristics of the unspecified neuro disease cohort

Table S6. Demographic characteristics of the unspecified respiratory disease cohort

Table S7. Classification performance metrics

Table S8. Large Language Models reference

Table S9. Multiclass classification results for infectious disease for all years

Table S10. Per-disease classification results for infectious disease for all years

Table S11. Ablation study for all infectious diseases

Table S12. Ablation study for the 15 rarest infectious diseases

Table S13. Multiclass unspecified disease classification performance for all years

Table S14. Number of training cases per unspecified disease for all years

Table S15. Number of evaluation cases per specified disease for all years

Table S16. Lead time and FDR for spatiotemporal detection

Table S17. SaTScan's configuration file used for the outbreak detection task

Figure S1. Class distribution for the infectious disease cohort

Figure S2. Class distribution for the unspecified disease cohort

*Table S1. List of 55 infectious disease considered in the classification problem and the associated ICD Codes. Diseases in the 15 rarest disease set used in the experiment (Table 2) in the manuscript are indicated with "*" in front of the disease name.*

| Infectious Disease | ICD Codes |
|---|---|
| **Anthrax, human or animal** | 022.0, 022.3, A22.7 |
| ***Babesiosis** | 088.82, B60.00, B60.02, B60.10, B60.11, B60.13, B60.19, B60.8 |
| **Botulism (Infant, Foodborne)** | 005.1, A05.1 |
| ***Brucellosis, human** | 023.0, 023.8, 023.9, A23.0, A23.1, A23.3, A23.8, A23.9 |
| **Campylobacteriosis** | 008.43, A04.5 |
| **Chickenpox (Varicella) (Outbreaks, hospitalizations and deaths)** | 052.0, 052.1, 052.2, 052.7, 052.8, 052.9, B01.0, B01.11, B01.2, B01.81, B01.89, B01.9 |
| **Chikungunya Virus Infection** | 065.4, A92.0 |
| **Cholera** | 001.0, 001.9, A00.0, A00.9 |
| **Ciguatera Fish Poisoning** | T61.01XA |
| ***Creutzfeldt-Jacob Disease (CJD) and other Transmissible spongiform** | 046.11, 046.19, A81.00, A81.01, A81.09 |
| ***Cryptosporidiosis** | 007.4, A07.2 |
| **Dengue Virus Infection** | 061, A90, A91 |
| **Diphtheria** | 032.0, 032.3, 032.81, 032.83, 032.84, 032.85, 032.89, 032.9, A36.0, A36.2, A36.3, A36.84, A36.89, A36.9 |
| **Ehrlichiosis** | 082.40, 082.41, 082.49, A77.40, A77.41, A77.49, A79.81 |
| ***Escherichia coli: Shiga toxin producing (STEC) including E.** | 041.41, 041.42, 041.43, B96.21, B96.22, B96.23 |
| **Haemophilus influenzae, invasive disease, all serotypes** | 041.5, 482.2, A49.2, B96.3, J14, J20.1 |
| **Hantavirus Infections** | 079.81, B33.4 |
| ***Hemolytic Uremic Syndrome** | 283.11, D59.3, D59.30, D59.31 |
| **Hepatitis A, acute infection** | 070.0, 070.1, B15.0, B15.9 |
| **Legionellosis** | 040.89, 482.84, A48.1, A48.2 |
| ***Listeriosis** | 027.0, A32.11, A32.12, A32.7, A32.89, A32.9 |
| **Malaria** | 084.0, 084.1, 084.4, 084.5, 084.6, 084.9, B50.0, B50.8, B50.9, B51.9, B52.0, B52.8, B52.9, B53.8, B54 |
| **Measles (Rubeola)** | 055.79, 055.8, 055.9, B05.2, B05.89, B05.9 |
| **Meningitis, Specify Etiology: Viral** | 047.0, 047.1, 047.8, 047.9, A87.0, A87.2, A87.8, A87.9 |
| ***Meningococcal Infections** | 036.0, 036.1, 036.2, 036.42, 036.81, 036.82, 036.89, 036.9, A39.0, A39.1, A39.2, A39.4, A39.81, A39.83, A39.9 |
| **Monkeypox or orthopox virus infection** | B04 |
| **Mosquito encephalitis** | 062.0, 062.2, 062.3, 062.9, A83.0, A83.2, A83.3, A83.5, A83.6, A83.8 |
| ***Mumps** | B26.0, B26.84, B26.9 |
| **Paratyphoid Fever** | 002.1, 002.2, 002.3, 002.9, A01.1, A01.3, A01.4 |
| **Pertussis (Whooping Cough)** | 033.0, 033.1, 033.9, A37.00, A37.01, A37.10, A37.11, A37.80, A37.90, A37.91 |
| **Plague, human or animal** | 020.2, 020.8, A20.0, A20.7, A20.8, A20.9 |

| | |
|---|---|
| ***Poliovirus Infection** | 045.00, 045.01, 045.10, 045.90, A80.30, A80.39, A80.9 |
| **Psittacosis** | 073.0, 073.8, 073.9, A70 |
| ***Q Fever** | 083.0, A78 |
| **Rabies, human or animal** | 071, A82.0, A82.1, A82.9 |
| ***Relapsing Fever** | 087.0, 087.1, 087.9, A68.0, A68.1, A68.9 |
| **Rickettsial Diseases (non-Rocky Mountain Spotted Fever), including Typhus and Typhus-like illnesses** | 065.3, 066.1, 082.9, 083.2, 083.8, 083.9, A75.2, A75.3, A77.8, A77.9, A79.1, A79.89, A79.9 |
| **Rocky Mountain Spotted Fever** | 082.0, A77.0 |
| **Rubella Syndrome, congenital** | B06.02, B06.89, B06.9, P35.0, Z20.4 |
| **Salmonellosis (Other than Typhoid)** | 003.0, 003.1, 003.20, 003.23, 003.29, 003.9, A02.0, A02.1, A02.20, A02.22, A02.23, A02.24, A02.25, A02.29, A02.8, A02.9 |
| **Scombroid Fish Poisoning** | T61.11XA, T61.14XA |
| **Shigellosis** | 004.0, 004.1, 004.2, 004.3, 004.8, 004.9, A03.0, A03.1, A03.2, A03.3, A03.8, A03.9 |
| **Syphilis (all stages, including congenital)** | 090.0, 090.2, 090.3, 090.40, 090.49, 090.5, 090.6, 090.7, 090.9, 091.0, 091.2, 091.3, 091.4, 091.51, 091.52, 091.82, 091.9, 092.0, 092.9, 093.0, 093.1, 093.22, 093.24, 093.89, 094.0, 094.1, 094.2, 094.3, 094.81, 094.82, 094.84, 094.85, 094.87, 094.89, 094.9, 095.1, 095.4, 095.5, 095.7, 095.8, 095.9, 096, 097.0, 097.1, 097.9, A50.02, A50.1, A50.2, A50.31, A50.40, A50.52, A50.53, A50.9, A51.0, A51.1, A51.2, A51.31, A51.32, A51.39, A51.43, A51.44, A51.49, A51.5, A51.9, A52.01, A52.02, A52.03, A52.10, A52.11, A52.13, A52.14, A52.15, A52.16, A52.17, A52.19, A52.2, A52.3, A52.71, A52.74, A52.76, A52.77, A52.79, A52.8, A52.9, A53.0, A53.9 |
| ***Tetanus** | 037, A33, A34, A35 |
| **Tick encephalitis** | A84.9 |
| ***Trichinosis** | 124, B75 |
| **Tuberculosis** | 010.00, 010.05, 010.06, 010.12, 010.13, 010.80, 010.84, 010.90, 010.92, 010.93, 011.00, 011.03, 011.10, 011.12, 011.15, 011.20, 011.21, 011.22, 011.23, 011.24, 011.25, 011.26, 011.33, 011.40, 011.41, 011.42, 011.44, 011.50, 011.51, 011.52, 011.53, 011.54, 011.56, 011.60, 011.62, 011.63, 011.64, 011.70, 011.80, 011.82, 011.83, 011.84, 011.86, 011.90, 011.91, 011.92, 011.93, 011.94, 011.95, 011.96, 012.00, 012.04, 012.10, 012.32, 012.80, 012.81, 012.83, 013.00, 013.04, 013.10, 013.13, 013.20, 013.30, 013.32, 013.40, 013.53, 013.54, 014.00, 014.03, 014.04, 014.06, 014.80, 014.82, 014.84, 014.86, 015.00, 015.02, 015.04, 015.10, 015.24, 015.50, 015.60, 015.66, 015.70, 015.75, 015.80, 015.90, 016.00, 016.10, 016.20, 016.30, 016.50, 016.60, 016.70, 016.90, 017.00, 017.06, 017.10, 017.20, 017.22, 017.23, 017.26, 017.30, 017.40, 017.60, 017.70, 017.90, 018.00, 018.03, 018.04, 018.05, 018.82, 018.90, 018.94, 018.95, 018.96, A15.0, A15.4, A15.5, A15.6, A15.7, A15.8, A15.9, A17.0, A17.1, A17.81, A17.82, A17.9, A18.01, A18.02, A18.03, A18.10, A18.11, A18.12, A18.13, A18.14, A18.15, A18.17, A18.18, A18.2, A18.31, A18.32, A18.39, A18.4, A18.50, A18.51, A18.52, A18.53, A18.54, A18.6, A18.83, A18.84, A19.0, A19.1, A19.2, A19.8, A19.9 |
| **Tularemia, human** | 021.0, 021.9, A21.2, A21.7, A21.8, A21.9 |
| **Typhoid Fever, Cases and Carriers** | 002.0, A01.00, A01.02, A01.03, A01.09 |
| **Vibrio Infections** | 005.4, 005.81, A05.3, A05.5 |
| **Viral Hemorrhagic Fevers, human or animal (e.g., Crimean-Congo, Ebola,** | A98.4, A98.5 |

| | |
|---|---|
| **Lassa, and Marburg viruses)** | |
| **West Nile Virus (WNV) Infection** | 066.40, 066.41, 066.42, 066.49, A92.30, A92.31, A92.32, A92.39 |
| **Yellow Fever** | A95.1 |
| **Yersiniosis** | 008.44, A04.6 |
| ***Zika Virus Infection** | A92.5 |

*Table S2. List of ICD Codes associated with unspecified GI, Neuro and Respiratory diseases.*

| Disease | ICD Codes |
|---|---|
| **Unspecified GI** | K52 - *Other And Unsp Noninfective Gastroenteritis And Colitis*,<br>K65 - *Peritonitis*,<br>K71 - *Toxic Liver Disease,*<br>K72 - *Hepatic Failure, Not Elsewhere Classified*,<br>K75 - *Other Inflammatory Liver Diseases*,<br>K85 - *Acute Pancreatitis.* |
| **Unspecified Neuro** | G02 - *Meningitis In Oth Infec/Parastc Diseases Classd Elswhr,*<br>G03 - *Meningitis Due To Other And Unspecified Causes*,<br>G04 - *Encephalitis, Myelitis And Encephalomyelitis*,<br>G05 - *Encphlts, Myelitis & Encephalomyelitis In Dis Classd Elswhr*,<br>G44 - *Other Headache Syndromes*,<br>G64 - *Other Disorders Of Peripheral Nervous System*,<br>G65 - *Sequelae Of Inflammatory And Toxic Polyneuropathies*,<br>G72 - *Other And Unspecified Myopathies*,<br>G92 - *Toxic Encephalopathy*,<br>G95 - *Other And Unspecified Diseases Of Spinal Cord.* |
| **Unspecified Respiratory** | J12 - *Viral Pneumonia, Not Elsewhere Classified*,<br>J15 - *Bacterial Pneumonia, Not Elsewhere Classified*,<br>J18 - *Pneumonia, Unspecified Organism*,<br>J22 - *Unspecified Acute Lower Respiratory Infection,*<br>J68 - *Resp Cond D/T Inhalation Of Chemicals, Gas, Fumes And Vapors*,<br>J80 - *Acute Respiratory Distress Syndrome*,<br>J81 - *Pulmonary Edema*,<br>J96 - *Respiratory Failure, Not Elsewhere Classified.* |

*Table S3. Demographic characteristics of the infectious disease cohort. (Race abbreviations: AP – Asian or Pacific Islanders, Unspecified, AS – Asian, BA – Black or African American, HP – Native Hawaiian or Other Pacific Islander, IN – American India or Alaska Native, MU – Multiracial, WH – White)*

| | **Patient** | **Control** |
|---|---|---|
| Number of individuals | 65,601 | 656,010 |
| Age (median [range]) | 37 [0-103] | 37 [0-107] |
| Male – no. (%) | 37,056 (56.49%) | 370,562 (56.49%) |
| **Race distribution** | | |
| AP – no. (%) | 36 (0.05%) | 382 (0.06%) |
| AS – no. (%) | 11,931 (18.19%) | 119,514 (18.22%) |
| BA – no. (%) | 6,394 (9.75%) | 56,819 (8.66%) |
| HP – no. (%) | 526 (0.80%) | 5,012 (0.76%) |
| IN – no. (%) | 309 (0.47%) | 2,966 (0.45%) |
| MU – no. (%) | 3,456 (5.27%) | 31,765 (4.84%) |
| WH – no. (%) | 25,335 (38.62%) | 276,312 (42.12%) |
| Unknown – no. (%) | 15,409 (23.49%) | 132,194 (20.15%) |
| Missing – no. (%) | 2,205 (3.36%) | 31,046 (4.73%) |
| Number of individuals without medical record | 96 (0.15%) | 65,625 (10.00%) |
| Median (first-to-last) visit interval (days) | 310 (0.47%) | 328 (0.05%) |

*Table S4. Demographic characteristics of the unspecified gastrointestinal (GI) cohort. (Race abbreviations: AP – Asian or Pacific Islanders, Unspecified, AS – Asian, BA – Black or African American, HP – Native Hawaiian or Other Pacific Islander, IN – American India or Alaska Native, MU – Multiracial, WH – White)*

| | **Patient** | **Control** |
|---|---|---|
| Number of individuals | 60,514 | 121,028 |
| Age (median [range]) | 44 [0-101] | 44 [0-102] |
| Male – no. (%) | 27,950 (46.19%) | 55,899 (46.19%) |
| **Race distribution** | | |
| AP – no. (%) | 47 (0.08%) | 95 (0.08%) |
| AS – no. (%) | 10,108 (18.19%) | 21,875 (18.07%) |
| BA – no. (%) | 4,721 (7.80%) | 9,938 (8.21%) |
| HP – no. (%) | 436 (0.72%) | 940 (0.78%) |
| IN – no. (%) | 316 (0.52%) | 570 (0.47%) |
| MU – no. (%) | 3,361 (5.55%) | 6,126 (5.06%) |
| WH – no. (%) | 25,706 (42.48%) | 53,641 (44.32%) |
| Unknown – no. (%) | 14,027 (23.18%) | 23,273 (19.23%) |
| Missing – no. (%) | 1,792 (2.96%) | 4,570 (3.78%) |
| Number of individuals without medical record | 369 (0.61%) | 1 (0.00%) |
| Median (first-to-last) visit interval (days) | 317 (0.52%) | 294 (0.24%) |

*Table S5. Demographic characteristics of the unspecified neuro cohort. (Race abbreviations: AP – Asian or Pacific Islanders, Unspecified, AS – Asian, BA – Black or African American, HP – Native Hawaiian or Other Pacific Islander, IN – American India or Alaska Native, MU – Multiracial, WH – White)*

| | Patient | Control |
|---|---|---|
| Number of individuals | 25,882 | 51,764 |
| Age (median [range]) | 45 [0-101] | 45 [0-100] |
| Male – no. (%) | 9,253 (35.75%) | 33,257 (64.25%) |
| **Race distribution** | | |
| AP – no. (%) | 15 (0.06%) | 16 (0.03%) |
| AS – no. (%) | 4,170 (16.11%) | 7,464 (14.42%) |
| BA – no. (%) | 2,597 (10.03%) | 5,900 (11.40%) |
| HP – no. (%) | 180 (0.70%) | 322 (0.62%) |
| IN – no. (%) | 119 (0.46%) | 299 (0.58%) |
| MU – no. (%) | 1,413 (5.46%) | 3,019 (5.83%) |
| WH – no. (%) | 10,682 (41.27%) | 24,740 (47.79%) |
| Unknown – no. (%) | 6,102 (23.58%) | 8,896 (17.19%) |
| Missing – no. (%) | 604 (2.33%) | 1,108 (2.14%) |
| Number of individuals without medical record | 79 (0.31%) | 71 (0.14%) |
| Median (first-to-last) visit interval (days) | 325 (1.26%) | 295 (0.57%) |

*Table S6. Demographic characteristics of the unspecified respiratory cohort. (Race abbreviations: AP – Asian or Pacific Islanders, Unspecified, AS – Asian, BA – Black or African American, HP – Native Hawaiian or Other Pacific Islander, IN – American India or Alaska Native, MU – Multiracial, WH – White)*

| | Patient | Control |
|---|---|---|
| Number of individuals | 37,982 | 75,964 |
| Age (median [range]) | 59 [0-104] | 59 [0-104] |
| Male – no. (%) | 18,608 (48.99%) | 37,217 (48.99%) |
| **Race distribution** | | |
| AP – no. (%) | 51 (0.13%) | 72 (0.09%) |
| AS – no. (%) | 5,560 (14.64%) | 10,903 (18.22%) |
| BA – no. (%) | 3,259 (8.58%) | 6,961 (9.16%) |
| HP – no. (%) | 302 (0.80%) | 391 (0.51%) |
| IN – no. (%) | 193 (0.51%) | 323 (0.43%) |
| MU – no. (%) | 2,366 (6.23%) | 4,536 (5.97%) |
| WH – no. (%) | 19,432 (15.67%) | 39,200 (51.60%) |
| Unknown – no. (%) | 5,951 (15.67%) | 11,544 (15.20%) |
| Missing – no. (%) | 868 (2.29%) | 2,034 (2.68%) |
| Number of individuals without medical record | 10 (0.03%) | 113 (0.15%) |
| Median (first-to-last) visit interval (days) | 335 (0.88%) | 302 (0.40%) |

*Table S7. Classification performance metrics.*

| Metric | Description | Mathematical Definition |
|---|---|---|
| **F1 Score (Micro and Macro)** | The F1 score balances precision (proportion of correct positive predictions) and recall (proportion of actual positives correctly identified) using their harmonic mean. Originally for binary classification, it extends to multi-class models in two key ways:<br><br>• **Micro F1 Score** aggregates true positives, false positives, and false negatives across all classes, providing an overall performance measure. It emphasizes larger classes, making it sensitive to class imbalance, common in infectious disease classification.<br><br>• **Macro F1 Score** computes F1 for each class individually and averages them equally, making it useful when all classes are important, even if some are underrepresented. | $F1 = 2 * \frac{Precision \, * \, Recall}{Precision \, + \, Recall}$ |
| **Balanced Accuracy** | In binary classification, Balanced Accuracy calculates the average of the recall for each of the two classes, ensuring that both minority and majority classes are equally considered. This metric mitigates the bias that can occur when one class dominates the dataset, providing a more holistic view of the model's performance across all classes. For the multilabel case, the Macro Balanced Accuracy is reported. | $BA = \frac{1}{2}\left(\frac{TP}{TP+FN} + \frac{TN}{TN+FP}\right)$ |
| **Matthew Correlation Coefficient (MCC)** | MCC considers true positives, true negatives, false positives, and false negatives in a balanced way. For binary classification, MCC ranges from -1 (perfect misclassification) to +1 (perfect classification), with 0 indicating random performance. For multiclass classification, perfect classification is still +1, but the minimum value depends of the number and distribution of ground true labels and it can be somewhere between -1 and 0. MCC captures correlations between predicted and true labels across all classes, making it particularly useful for multilabel tasks with imbalanced datasets. | $MCC = \frac{c * s - \sum_k^K p_k * t_k}{\sqrt{(s^2 - \sum_k^K p_k^2) * (s^2 - \sum_k^K t_k^2)}}$<br><br>• $t_k = \sum_i^K C_{ik}$ is the number of times class $k$ truly occurred;<br>• $p_k = \sum_i^K C_{ki}$ is the number of times class $k$ was predicted;<br>• $c = \sum_k^K C_{kk}$ is the total number of samples correctly predicted; and<br>• $s = \sum_i^K \sum_j^K C_{ij}$ is the total number of samples. |

*Table S8. Large Language Models reference.*

| Model Name | Pre-Trained Model Reference |
|---|---|
| **BERT Base** | https://huggingface.co/google-bert/bert-base-uncased |
| **BioBERT** | https://huggingface.co/dmis-lab/biobert-v1.1 |
| **BioClinicalBERT** | https://huggingface.co/emilyalsentzer/Bio_ClinicalBERT |

*Table S9. Multiclass classification performance metrics of XGBoost with count-based featurization and BioclinicalBERT using raw medical events for the infectious disease classification over the years. For each evaluation year, the model was trained on medical records up to the previous year. For example, for the 2014 evaluation year, the models were trained on all available data up to 2013. Both models were trained with sample weighted strategy.*

| | | 2014 | 2015 | 2016 | 2017 | 2018 | 2019 | 2020 | 2021 | 2022 |
|---|---|---|---|---|---|---|---|---|---|---|
| **F1-Micro** | XGBoost | 0.82 | 0.78 | 0.77 | 0.77 | 0.78 | 0.78 | 0.79 | 0.80 | 0.76 |
| | BioClinicalBERT | 0.81 | 0.78 | 0.77 | 0.78 | 0.80 | 0.77 | 0.77 | 0.81 | 0.78 |
| **F1-Macro** | XGBoost | 0.38 | 0.37 | 0.38 | 0.36 | 0.38 | 0.39 | 0.39 | 0.40 | 0.36 |
| | BioClinicalBERT | 0.35 | 0.35 | 0.37 | 0.39 | 0.39 | 0.34 | 0.33 | 0.37 | 0.32 |
| **MCC** | XGBoost | 0.67 | 0.61 | 0.60 | 0.60 | 0.62 | 0.61 | 0.62 | 0.63 | 0.57 |
| | BioClinicalBERT | 0.68 | 0.62 | 0.61 | 0.62 | 0.66 | 0.62 | 0.61 | 0.66 | 0.61 |
| **BA-Macro** | XGBoost | 0.64 | 0.66 | 0.66 | 0.66 | 0.68 | 0.67 | 0.68 | 0.71 | 0.69 |
| | BioClinicalBERT | 0.67 | 0.67 | 0.67 | 0.67 | 0.69 | 0.69 | 0.69 | 0.68 | 0.70 |

*Table S10. Average F1-Score of the model's performance per disease, comparing Unweighted and Weighted loss strategies. Metrics are scaled from 0 to 1, with 1 indicating perfect prediction accuracy. Missing values (dashes) signify cases where the F1-score could not be computed due to the absence of positive predictions or true positives.*

| | **Unweighted** | | | | **Weighted** | | | |
|---|---|---|---|---|---|---|---|---|
| | XGBoost | BERT | BioBERT | Bio-ClinicalBERT | XGBoost | BERT | BioBERT | Bio-ClinicalBERT |
| **Control (131,202)** | 0.91 | 0.93 | 0.93 | 0.94 | 0.90 | 0.91 | 0.91 | 0.92 |
| **Syphilis (17,748)** | 0.74 | 0.75 | 0.76 | 0.77 | 0.74 | 0.74 | 0.73 | 0.76 |
| **Campylobacteriosis (8,445)** | 0.59 | 0.62 | 0.61 | 0.61 | 0.58 | 0.60 | 0.58 | 0.62 |
| **Chickenpox (6,671)** | 0.56 | 0.58 | 0.59 | 0.60 | 0.54 | 0.55 | 0.53 | 0.55 |
| **Pertussis (6,131)** | 0.57 | 0.62 | 0.62 | 0.66 | 0.50 | 0.58 | 0.57 | 0.59 |
| **Tuberculosis (6,060)** | 0.54 | 0.47 | 0.45 | 0.50 | 0.55 | 0.44 | 0.45 | 0.48 |
| **Salmonellosis (5,279)** | 0.42 | 0.40 | 0.40 | 0.46 | 0.42 | 0.38 | 0.36 | 0.42 |
| **Shigellosis (3,474)** | 0.25 | 0.22 | 0.28 | 0.14 | 0.32 | 0.28 | 0.31 | 0.30 |
| **Meningitis (2,872)** | 0.58 | 0.33 | 0.34 | 0.42 | 0.53 | 0.30 | 0.35 | 0.40 |
| **Haemophilus (1,624)** | 0.38 | 0.29 | 0.28 | 0.36 | 0.35 | 0.27 | 0.26 | 0.31 |

| | | | | | | | | |
|---|---|---|---|---|---|---|---|---|
| **Hepatitis A (1,239)** | 0.09 | 0.10 | 0.11 | 0.16 | 0.19 | 0.12 | 0.15 | 0.14 |
| **Dengue Virus Infection (480)** | 0.18 | 0.30 | 0.28 | 0.32 | 0.21 | 0.25 | 0.27 | 0.31 |
| **Hemolytic Uremic Synd. (385)** | 0.18 | 0.10 | 0.21 | 0.10 | 0.23 | 0.17 | 0.20 | 0.16 |
| **Legionelllosis (363)** | 0.26 | | 0.06 | 0.21 | 0.22 | 0.12 | 0.16 | 0.24 |
| **Malaria (354)** | 0.38 | 0.43 | 0.40 | 0.43 | 0.46 | 0.38 | 0.38 | 0.42 |
| **West Nile Virus (324)** | 0.14 | 0.20 | 0.13 | 0.15 | 0.17 | 0.10 | 0.17 | 0.22 |
| **E. coli (317)** | - | - | - | - | 0.09 | 0.08 | 0.06 | 0.11 |
| **Typhoid Fever (282)** | 0.11 | - | 0.11 | 0.11 | 0.19 | 0.08 | 0.09 | 0.14 |
| **Relapsing Fever (278)** | 0.17 | 0.19 | 0.18 | 0.16 | 0.24 | 0.12 | 0.10 | 0.14 |
| **Cryptosporidiosis (248)** | 0.55 | 0.60 | 0.46 | 0.65 | 0.58 | 0.51 | 0.51 | 0.56 |
| **Q Fever (221)** | 0.67 | 0.46 | 0.53 | 0.62 | 0.63 | 0.51 | 0.39 | 0.68 |
| **Poliovirus Infection (195)** | 0.25 | - | - | - | 0.32 | 0.07 | 0.07 | 0.20 |
| **Rubella Synd. (174)** | 0.09 | - | - | - | 0.25 | 0.12 | 0.18 | 0.15 |
| **Trichinosis (172)** | 0.22 | 0.42 | 0.52 | 0.51 | 0.33 | 0.45 | 0.40 | 0.44 |
| **Mumps (167)** | 0.11 | 0.35 | 0.25 | 0.28 | 0.13 | 0.22 | 0.15 | 0.18 |
| **Tetanus (142)** | 0.33 | 0.18 | - | 0.31 | 0.21 | 0.14 | 0.08 | 0.13 |
| **Listeriosis (135)** | - | - | - | - | 0.11 | 0.07 | 0.08 | 0.07 |
| **Creutzfeldt-Jacob Disease (106)** | 0.34 | - | - | - | 0.38 | - | 0.09 | 0.12 |
| **Rickettsial Diseases (99)** | - | - | - | - | 0.08 | - | 0.08 | - |
| **Chikungunya Virus Infection (97)** | - | - | - | - | - | 0.08 | - | - |
| **Babesiosis (92)** | - | - | - | - | - | 0.19 | 0.10 | 0.15 |
| **Brucellosis (76)** | 0.18 | - | - | - | - | 0.20 | 0.34 | 0.28 |
| **Zika Virus Infection (75)** | 0.10 | - | - | - | 0.06 | 0.04 | 0.08 | 0.10 |
| **Botulism (65)** | 0.70 | - | - | - | 0.77 | 0.23 | 0.24 | 0.32 |
| **Vibrio Infections (65)** | - | - | - | - | - | 0.11 | 0.10 | 0.12 |
| **Rabies (56)** | 0.91 | - | - | - | - | 0.13 | 0.08 | 0.30 |
| **Scombroid Fish Poisoning (43)** | 0.35 | - | - | - | 0.44 | 0.23 | 0.40 | 0.35 |

*Table S11. Average multiclass performance of the BioClinicalBERT model (with sample weighting) considering all diseases from 2019 to 2022 evaluated across different combinations of medical event types. Patient demographic information was included in all ablation study experiments. Among individual modalities, medication was the most informative, while the combination of all modalities yielded the best overall performance.*

| Data Types | F1-Micro | F1-Macro | MCC | BA-Macro |
|---|---|---|---|---|
| **Vitals** | 0.66 | 0.28 | 0.30 | 0.53 |
| **Diagnosis** | 0.64 | 0.22 | 0.40 | 0.60 |
| **Labs** | 0.68 | 0.27 | 0.45 | 0.60 |
| **Medications** | 0.69 | 0.28 | 0.46 | 0.63 |
| **Labs + Diagnosis** | 0.68 | 0.27 | 0.47 | 0.62 |
| **Medications + Diagnosis** | 0.72 | 0.30 | 0.53 | 0.67 |
| **Medications + Labs** | 0.74 | 0.34 | 0.56 | 0.68 |
| **Medications + Labs + Diagnosis** | 0.76 | 0.34 | 0.60 | 0.67 |
| **All** | **0.78** | **0.34** | **0.62** | **0.69** |

*Table S12. Average multiclass performance of the BioClinicalBERT model (with sample weighting) on the 15 rarest diseases, over the period of 2019 through 2022, evaluated across different combinations of medical event types. Classification performance only considering the rarest infectious diseases, medications and laboratory tests only, aside from the patient demographics information, yielded the best results for all metrics.*

| Modalities | F1-Micro | F1-Macro | MCC | BA-Macro |
|---|---|---|---|---|
| **Vitals** | 0.10 | 0.32 | 0.06 | 0.50 |
| **Diagnosis** | 0.21 | 0.28 | 0.18 | 0.57 |
| **Labs** | 0.22 | 0.32 | 0.20 | 0.57 |
| **Medications** | 0.34 | 0.41 | 0.31 | 0.61 |
| **Labs + Diagnosis** | 0.26 | 0.32 | 0.24 | 0.59 |
| **Medications + Diagnosis** | 0.37 | 0.41 | 0.35 | 0.65 |
| **Medications + Labs** | **0.39** | **0.45** | **0.36** | **0.65** |
| **Medications + Labs + Diagnosis** | 0.39 | 0.42 | 0.36 | 0.63 |
| **All** | 0.39 | 0.42 | 0.35 | 0.64 |

*Table S13. Multiclass classification performance for the Unspecified Disease classification problems over all years of available data. Results show a consistent improved performance of BioclinicalBERT model over XGBoost.*

| Year | Model | BA-Macro | F1-Macro | F1-Micro | MCC |
|---|---|---|---|---|---|
| **2014** | XGBoost | 0.73 | 0.53 | 0.72 | 0.46 |
| | BioClinicalBERT | 0.76 | 0.57 | 0.75 | 0.53 |
| **2015** | XGBoost | 0.73 | 0.54 | 0.73 | 0.48 |
| | BioClinicalBERT | 0.77 | 0.58 | 0.76 | 0.55 |
| **2016** | XGBoost | 0.73 | 0.53 | 0.72 | 0.47 |
| | BioClinicalBERT | 0.76 | 0.57 | 0.75 | 0.53 |
| **2017** | XGBoost | 0.73 | 0.53 | 0.72 | 0.47 |
| | BioClinicalBERT | 0.77 | 0.57 | 0.75 | 0.53 |
| **2018** | XGBoost | 0.72 | 0.52 | 0.72 | 0.45 |
| | BioClinicalBERT | 0.74 | 0.51 | 0.71 | 0.48 |
| **2019** | XGBoost | 0.72 | 0.52 | 0.72 | 0.46 |
| | BioClinicalBERT | 0.76 | 0.58 | 0.76 | 0.54 |
| **2020** | XGBoost | 0.70 | 0.50 | 0.72 | 0.45 |
| | BioClinicalBERT | 0.74 | 0.56 | 0.75 | 0.51 |
| **2021** | XGBoost | 0.70 | 0.51 | 0.72 | 0.44 |
| | BioClinicalBERT | 0.74 | 0.56 | 0.76 | 0.52 |
| **2022** | XGBoost | 0.72 | 0.52 | 0.72 | 0.46 |
| | BioClinicalBERT | 0.75 | 0.55 | 0.73 | 0.51 |

*Table S14. Number of training samples per class for each year for the Unspecified Disease classification problem.*

| Period | Control | Known Disease | Unsp. GI | Unsp. Neuro | Unsp. Respiratory | Total |
|---|---|---|---|---|---|---|
| **2010 – 2013** | 116,349 | 30,908 | 13,041 | 8,744 | 6,964 | 176,006 |
| **2010 – 2014** | 150,071 | 39,032 | 17,716 | 8,997 | 11,262 | 227,078 |
| **2010 – 2015** | 193,497 | 51,189 | 22,673 | 10,903 | 14,512 | 292,774 |
| **2010 – 2016** | 228,937 | 59,717 | 27,536 | 12,900 | 17,340 | 346,430 |
| **2010 – 2017** | 265,008 | 68,467 | 32,533 | 14,756 | 20,311 | 401,075 |
| **2010 – 2018** | 300,125 | 77,135 | 37,445 | 16,605 | 23,018 | 454,328 |
| **2010 – 2019** | 350,918 | 93,571 | 42,446 | 18,335 | 26,097 | 531,367 |
| **2010 – 2020** | 379,380 | 100,740 | 45,769 | 19,622 | 28,994 | 574,445 |
| **2010 – 2021** | 410,410 | 108,472 | 49,862 | 21,100 | 31,714 | 621,558 |

*Table S15. Number of evaluation cases for each class per year for the Unspecified Disease classification problem.*

| Year | Control | Known Disease | Unsp GI | Unsp Neuro | Unsp Respiratory | Total |
|---|---|---|---|---|---|---|
| **2014** | 36,704 | 9,601 | 4,675 | 2,033 | 2,518 | 55,531 |
| **2015** | 40,555 | 10,686 | 4,957 | 1,906 | 3,250 | 61,354 |
| **2016** | 38,713 | 10,235 | 4,863 | 1,997 | 2,828 | 58,636 |
| **2017** | 39,394 | 10,500 | 4,997 | 1,856 | 2,971 | 59,718 |
| **2018** | 38,525 | 10,403 | 4,912 | 1,849 | 2,707 | 55,689 |
| **2019** | 39,156 | 10,367 | 5,001 | 1,730 | 3,079 | 59,333 |
| **2020** | 29,817 | 7,886 | 3,323 | 1,287 | 2,847 | 45,160 |
| **2021** | 32,487 | 8,505 | 4,093 | 1,478 | 2,770 | 49,333 |
| **2022** | 32,081 | 8,961 | 4,305 | 1,595 | 2,587 | 49,529 |

*Table S16. Average lead time and false detection rate for spatiotemporal outbreak detection using predicted case diagnoses combined with confirmed case diagnoses, as compared to outbreak detection using confirmed case diagnoses alone.*

| | XGBoost | | | BioClinicalBERT | | |
|---|---|---|---|---|---|---|
| **Infectious Disease (No. of diagnosed cases from 2010 to 2022)** | **Number of outbreaks detected with positive lead time/Number of true outbreaks** | **Average positive lead time (days)** | **Number of false detections** | **Number of outbreaks detected with positive lead time/Number of true outbreaks** | **Average positive lead time (days)** | **Number of false detections** |
| **Infectious Diseases** | | | | | | |
| Syphilis (17,748) | 17/20 | 23.8 | 4 | 15/20 | 24.5 | 3 |
| Campylobacteriosis (8,445) | 2/8 | 12.0 | 1 | 1/8 | 11.0 | 1 |
| Pertussis (6,131) | 3/8 | 7.3 | 1 | 2/8 | 5.0 | 1 |
| Tuberculosis (6,060) | 5/7 | 11.2 | 1 | 4/7 | 9.8 | 1 |
| Salmonellosis (5,279) | 2/7 | 5.5 | 1 | 1/7 | 4.0 | 1 |
| Shigellosis (3,474) | 2/8 | 2.5 | 1 | 3/8 | 9.0 | 1 |
| West Nile Virus (324) | 2/5 | 9.5 | 1 | 1/5 | 1.0 | 1 |
| Rubella Synd. (174) | 0/7 | — | 0 | 0/7 | — | 0 |
| Mumps (167) | 0/5 | — | 0 | 0/5 | — | 0 |
| **Unspecified Diseases** | | | | | | |
| Unspecified GI disease (41,146) | 0/11 | — | 1 | 2/11 | 1.0 | 1 |
| Unspecified respiratory disease (25,557) | 0/13 | — | 1 | 2/13 | 1.5 | 1 |

*Table S17. SaTScan's configuration file used for the outbreak detection task.*

| SatScan Configuration File |
|---|
| [Input]<br>;case data filename<br>CaseFile=observed_and_predicted_cases.cas<br>;source type (CSV=0, DBASE=1, SHAPE=2)<br>CaseFile-SourceType=0<br>;source field map (comma separated list of integers, oneCount, generatedId, shapeX, shapeY)<br>CaseFile-SourceFieldMap=1,2,3<br>;csv source delimiter (leave empty for space or tab delimiter) |

```
CaseFile-SourceDelimiter=" "
;csv source group character
CaseFile-SourceGrouper="
;csv source skip initial lines (i.e. meta data)
CaseFile-SourceSkip=0
;csv source first row column header
CaseFile-SourceFirstRowHeader=n
;control data filename
ControlFile=
;time precision (0=None, 1=Year, 2=Month, 3=Day, 4=Generic)
PrecisionCaseTimes=3
;study period start date (YYYY/MM/DD)
StartDate=2011/1/1
;study period end date (YYYY/MM/DD)
EndDate=2015/01/01
;population data filename
PopulationFile=census_tracts.pop
;source type (CSV=0, DBASE=1, SHAPE=2)
PopulationFile-SourceType=0
;source field map (comma separated list of integers, oneCount, generatedId, shapeX, shapeY)
PopulationFile-SourceFieldMap=1,2,3
;csv source delimiter (leave empty for space or tab delimiter)
PopulationFile-SourceDelimiter=" "
;csv source group character
PopulationFile-SourceGrouper="
;csv source skip initial lines (i.e. meta data)
PopulationFile-SourceSkip=0
;csv source first row column header
PopulationFile-SourceFirstRowHeader=n
;coordinate data filename
CoordinatesFile=census_tracts.geo
;source type (CSV=0, DBASE=1, SHAPE=2)
CoordinatesFile-SourceType=0
;source field map (comma separated list of integers, oneCount, generatedId, shapeX, shapeY)
CoordinatesFile-SourceFieldMap=1,2,3
;csv source delimiter (leave empty for space or tab delimiter)
CoordinatesFile-SourceDelimiter=" "
;csv source group character
CoordinatesFile-SourceGrouper="
;csv source skip initial lines (i.e. meta data)
CoordinatesFile-SourceSkip=0
;csv source first row column header
CoordinatesFile-SourceFirstRowHeader=n
;use grid file? (y/n)
UseGridFile=n
;grid data filename
GridFile=
;coordinate type (0=Cartesian, 1=latitude/longitude)
CoordinatesType=1

[Analysis]
;analysis type (1=Purely Spatial, 2=Purely Temporal, 3=Retrospective Space-Time, 4=Prospective Space-Time, 5=Spatial Variation in Temporal Trends, 6=Prospective Purely Temporal, 7=Seasonal Temporal)
AnalysisType=4
;model type (0=Discrete Poisson, 1=Bernoulli, 2=Space-Time Permutation, 3=Ordinal, 4=Exponential, 5=Normal, 6=Continuous Poisson, 7=Multinomial, 8=Rank, 9=UniformTime)
ModelType=0
;scan areas (1=High Rates(Poison,Bernoulli,STP); High Values(Ordinal,Normal); Short Survival(Exponential); Higher Trend(Poisson-SVTT), 2=Low Rates(Poison,Bernoulli,STP); Low Values(Ordinal,Normal); Long Survival(Exponential); Lower Trend(Poisson-SVTT), 3=Both Areas)
ScanAreas=1
;time aggregation units (0=None, 1=Year, 2=Month, 3=Day, 4=Generic)
```

```
TimeAggregationUnits=3
;time aggregation length (Positive Integer)
TimeAggregationLength=7

[Output]
;analysis main results output filename
ResultsFile=outbreaks.txt
;output Google Earth KML file (y/n)
OutputGoogleEarthKML=n
;output shapefiles (y/n)
OutputShapefiles=n
;output cartesian graph file (y/n)
OutputCartesianGraph=n
;output cluster information in ASCII format? (y/n)
MostLikelyClusterEachCentroidASCII=n
;output cluster information in dBase format? (y/n)
MostLikelyClusterEachCentroidDBase=n
;output cluster case information in ASCII format? (y/n)
MostLikelyClusterCaseInfoEachCentroidASCII=n
;output cluster case information in dBase format? (y/n)
MostLikelyClusterCaseInfoEachCentroidDBase=n
;output location information in ASCII format? (y/n)
CensusAreasReportedClustersASCII=n
;output location information in dBase format? (y/n)
CensusAreasReportedClustersDBase=n
;output risk estimates in ASCII format? (y/n)
IncludeRelativeRisksCensusAreasASCII=n
;output risk estimates in dBase format? (y/n)
IncludeRelativeRisksCensusAreasDBase=n
;output simulated log likelihoods ratios in ASCII format? (y/n)
SaveSimLLRsASCII=n
;output simulated log likelihoods ratios in dBase format? (y/n)
SaveSimLLRsDBase=n
;generate Google Maps output (y/n)
OutputGoogleMaps=n

[Multiple Data Sets]
; multiple data sets purpose type (0=Multivariate, 1=Adjustment)
MultipleDataSetsPurposeType=0

[Data Checking]
;study period data check (0=Strict Bounds, 1=Relaxed Bounds)
StudyPeriodCheckType=1
;geographical coordinates data check (0=Strict Coordinates, 1=Relaxed Coordinates)
GeographicalCoordinatesCheckType=0

[Locations Network]
;locations network filename
LocationsNetworkFilename=
;use locations network file
UseLocationsNetworkFile=n
;purpose of locations network file (0=Coordinates File Override, 1=Network Definition)
PurposeLocationsNetworkFile=1

[Line List]
;whether case file contains line list data (positive integer (y/n)
LineListCaseFile=n
;whether case file contains line list header row (positive integer (y/n)
LineListHeaderCaseFile=n
;file to store events ids
LineListEventCache=
;indication whether to include linelist events in kml output, grouped (y/n)
```

```
EventGroupKML=n
;label of line-list colummn that should be used to group events in KML file
EventGroupByKML=

[Spatial Neighbors]
;use neighbors file (y/n)
UseNeighborsFile=n
;neighbors file
NeighborsFilename=
;use meta locations file (y/n)
UseMetaLocationsFile=n
;meta locations file
MetaLocationsFilename=
;multiple coordinates type (0=OnePerLocation, 1=AtLeastOneLocation, 2=AllLocations)
MultipleCoordinatesType=0

[Spatial Window]
;maximum spatial size in population at risk (<=50%)
MaxSpatialSizeInPopulationAtRisk=50
;restrict maximum spatial size - max circle file? (y/n)
UseMaxCirclePopulationFileOption=n
;maximum spatial size in max circle population file (<=50%)
MaxSpatialSizeInMaxCirclePopulationFile=50
;maximum circle size filename
MaxCirclePopulationFile=
;restrict maximum spatial size - distance? (y/n)
UseDistanceFromCenterOption=n
;maximum spatial size in distance from center (positive integer)
MaxSpatialSizeInDistanceFromCenter=1
;include purely temporal clusters? (y/n)
IncludePurelyTemporal=n
;window shape (0=Circular, 1=Elliptic)
SpatialWindowShapeType=0
;elliptic non-compactness penalty (0=NoPenalty, 1=MediumPenalty, 2=StrongPenalty)
NonCompactnessPenalty=1
;isotonic scan (0=Standard, 1=Monotone)
IsotonicScan=0

[Temporal Window]
;minimum temporal cluster size (in time aggregation units)
MinimumTemporalClusterSize=1
;how max temporal size should be interpretted (0=Percentage, 1=Time)
MaxTemporalSizeInterpretation=1
;maximum temporal cluster size (<=90%)
MaxTemporalSize=28
;include purely spatial clusters? (y/n)
IncludePurelySpatial=n
;temporal clusters evaluated (0=All, 1=Alive, 2=Flexible Window)
IncludeClusters=0
;flexible temporal window start range (YYYY/MM/DD,YYYY/MM/DD)
IntervalStartRange=2000/1/1,2000/12/31
;flexible temporal window end range (YYYY/MM/DD,YYYY/MM/DD)
IntervalEndRange=2000/1/1,2000/12/31

[Cluster Restrictions]
;risk limit high clusters (y/n)
RiskLimitHighClusters=n
;risk threshold high clusters (1.0 or greater)
RiskThresholdHighClusters=1
;risk limit low clusters (y/n)
RiskLimitLowClusters=n
;risk threshold low clusters (0.000 - 1.000)
```

```
RiskThresholdLowClusters=1
;minimum cases in low rate clusters (positive integer)
MinimumCasesInLowRateClusters=0
;minimum cases in high clusters (positive integer)
MinimumCasesInHighRateClusters=2

[Space and Time Adjustments]
;time trend adjustment type (0=None, 2=LogLinearPercentage, 3=CalculatedLogLinearPercentage,
4=TimeStratifiedRandomization, 5=CalculatedQuadratic, 1=TemporalNonparametric)
TimeTrendAdjustmentType=0
;time trend adjustment percentage (>-100)
TimeTrendPercentage=0
;time trend type - SVTT only (Linear=0, Quadratic=1)
TimeTrendType=0
;adjust for weekly trends, nonparametric
AdjustForWeeklyTrends=n
;spatial adjustments type (0=None, 1=SpatiallyStratifiedRandomization, 2=SpatialNonparametric)
SpatialAdjustmentType=0
;use adjustments by known relative risks file? (y/n)
UseAdjustmentsByRRFile=n
;adjustments by known relative risks file name (with HA Randomization=1)
AdjustmentsByKnownRelativeRisksFilename=

[Inference]
;p-value reporting type (Default p-value=0, Standard Monte Carlo=1, Early Termination=2, Gumbel p-value=3)
PValueReportType=0
;early termination threshold
EarlyTerminationThreshold=50
;report Gumbel p-values (y/n)
ReportGumbel=n
;Monte Carlo replications (0, 9, 999, n999)
MonteCarloReps=999
;adjust for earlier analyses(prospective analyses only)? (y/n)
AdjustForEarlierAnalyses=n
;prospective surveillance start date (YYYY/MM/DD)
ProspectiveStartDate=2000/12/31
;perform iterative scans? (y/n)
IterativeScan=n
;maximum iterations for iterative scan (0-32000)
IterativeScanMaxIterations=10
;max p-value for iterative scan before cutoff (0.000-1.000)
IterativeScanMaxPValue=0.05

[Cluster Drilldown]
;perform detected cluster standard drilldown (y/n)
PerformStandardDrilldown=n
;perform detected cluster Bernoulli drilldown (y/n)
PerformBernoulliDrilldown=n
;minimum number of locations in detected cluster to perform drilldown (positive integer)
DrilldownMinimumClusterLocations=2
;minimum number of cases in detected cluster to perform drilldown (positive integer)
DrilldownMinimumClusterCases=10
;p-value cutoff of detected cluster to perform drilldown (0.000-1.000)
DrilldownClusterPvalueCutoff=0.05
;adjust for weekly trends, purely spatial Bernoulli drilldown
DrilldownAdjustForWeeklyTrends=n

[Miscellaneous Analysis]
;calculate Oliveira's F
CalculateOliveira=n
;number of bootstrap replications for Oliveira calculation (minimum=100, multiple of 100)
NumBootstrapReplications=1000
```

```
;p-value cutoff for cluster's in Oliveira calculation (0.000-1.000)
OliveiraPvalueCutoff=0.05
;frequency of prospective analyses type (0=Same Time Aggregation, 1=Daily, 2=Weekly, 3=Monthy, 4=Quarterly, 5=Yearly)
ProspectiveFrequencyType=1
;frequency of prospective analyses  (positive integer)
ProspectiveFrequency=1

[Power Evaluation]
;perform power evaluation - Poisson only (y/n)
PerformPowerEvaluation=n
;power evaluation method (0=Analysis And Power Evaluation Together, 1=Only Power Evaluation With Case File, 2=Only Power Evaluation With Defined Total Cases)
PowerEvaluationsMethod=0
;total cases in power evaluation
PowerEvaluationTotalCases=600
;critical value type (0=Monte Carlo, 1=Gumbel, 2=User Specified Values)
CriticalValueType=0
;power evaluation critical value .05 (> 0)
CriticalValue05=0
;power evaluation critical value .001 (> 0)
CriticalValue01=0
;power evaluation critical value .001 (> 0)
CriticalValue001=0
;power estimation type (0=Monte Carlo, 1=Gumbel)
PowerEstimationType=0
;number of replications in power step
NumberPowerReplications=1000
;power evaluation alternative hypothesis filename
AlternativeHypothesisFilename=
;power evaluation simulation method for power step (0=Null Randomization, 1=N/A, 2=File Import)
PowerEvaluationsSimulationMethod=0
;power evaluation simulation data source filename
PowerEvaluationsSimulationSourceFilename=
;report power evaluation randomization data from power step (y/n)
ReportPowerEvaluationSimulationData=n
;power evaluation simulation data output filename
PowerEvaluationsSimulationOutputFilename=

[Spatial Output]
;automatically launch map viewer - gui only (y/n)
LaunchMapViewer=y
;create compressed KMZ file instead of KML file (y/n)
CompressKMLtoKMZ=n
;whether to include cluster locations kml output (y/n)
IncludeClusterLocationsKML=y
;threshold for generating separate kml files for cluster locations (positive integer)
ThresholdLocationsSeparateKML=1000
;report hierarchical clusters (y/n)
ReportHierarchicalClusters=y
;criteria for reporting secondary clusters(0=NoGeoOverlap, 1=NoCentersInOther, 2=NoCentersInMostLikely, 3=NoCentersInLessLikely, 4=NoPairsCentersEachOther, 5=NoRestrictions)
CriteriaForReportingSecondaryClusters=0
;report gini clusters (y/n)
ReportGiniClusters=n
;gini index cluster reporting type (0=optimal index only, 1=all values)
GiniIndexClusterReportingType=0
;spatial window maxima stops (comma separated decimal values[<=50%] )
SpatialMaxima=1,2,3,4,5,6,8,10,12,15,20,25,30,40,50
;max p-value for clusters used in calculation of index based coefficients (0.000-1.000)
GiniIndexClustersPValueCutOff=0.05
;report gini index coefficents to results file (y/n)
ReportGiniIndexCoefficents=n
```

```
;restrict reported clusters to maximum geographical cluster size? (y/n)
UseReportOnlySmallerClusters=n
;maximum reported spatial size in population at risk (<=50%)
MaxSpatialSizeInPopulationAtRisk_Reported=50
;restrict maximum reported spatial size - max circle file? (y/n)
UseMaxCirclePopulationFileOption_Reported=n
;maximum reported spatial size in max circle population file (<=50%)
MaxSizeInMaxCirclePopulationFile_Reported=50
;restrict maximum reported spatial size - distance? (y/n)
UseDistanceFromCenterOption_Reported=n
;maximum reported spatial size in distance from center (positive integer)
MaxSpatialSizeInDistanceFromCenter_Reported=1

[Temporal Output]
;output temporal graph HTML file (y/n)
OutputTemporalGraphHTML=n
;temporal graph cluster reporting type (0=Only most likely cluster, 1=X most likely clusters, 2=Only significant clusters)
TemporalGraphReportType=0
;number of most likely clusters to report in temporal graph (positive integer)
TemporalGraphMostMLC=1
;significant clusters p-value cutoff to report in temporal graph (0.000-1.000)
TemporalGraphSignificanceCutoff=0.05

[Other Output]
;cluster significance by recurrence interval  (y/n)
ClusterSignificanceByRecurrence=n
;cluster significance recurrence interval cutoff (positive integer)
ClusterSignificanceRecurrenceCutoff=100
;cluster significance recurrence interval type (YEAR=1, DAY=3)
ClusterSignificanceRecurrenceCutoffType=4
;cluster significance by  p-value (y/n)
ClusterSignificanceByPvalue=n
;cluster significance p-value cutoff (0.000-1.000)
ClusterSignificancePvalueCutoff=0.05
;report critical values for .01 and .05? (y/n)
CriticalValue=n
;report cluster rank (y/n)
ReportClusterRank=n
;print ascii headers in output files (y/n)
PrintAsciiColumnHeaders=n
;user-defined title for results file
ResultsTitle=

[Email Alerts]
;whether to email user an analysis results summary
EmailResultsSummary=n
;list of users which are always emailed
EmailAlwaysRecipients=
;list of users which are emailed for significant events
EmailSignificantRecipients=
;subject line of email - no significant clusters
EmailSubjectLineNoSignificant=
;email message body - no significant clusters
EmailMessageBodyNoSignificant=
;subject line of email - significant clusters
EmailSubjectLineSignificant=
;email message body - significant clusters
EmailMessageBodySignificant=
;email message - attach results
EmailAttachResults=n

[Elliptic Scan]
```

```
;elliptic shapes - one value for each ellipse (comma separated decimal values)
EllipseShapes=1.5,2,3,4,5
;elliptic angles - one value for each ellipse (comma separated integer values)
EllipseAngles=4,6,9,12,15

[Power Simulations]
;simulation methods (0=Null Randomization, 1=N/A, 2=File Import)
SimulatedDataMethodType=0
;simulation data input file name (with File Import=2)
SimulatedDataInputFilename=
;print simulation data to file? (y/n)
PrintSimulatedDataToFile=n
;simulation data output filename
SimulatedDataOutputFilename=

[Run Options]
;number of parallel processes to execute (0=All Processors, x=At Most X Processors)
NumberParallelProcesses=0
;suppressing warnings? (y/n)
SuppressWarnings=n
;log analysis run to history file? (y/n)
LogRunToHistoryFile=n
;analysis execution method  (0=Automatic, 1=Successively, 2=Centrically)
ExecutionType=0

[System]
;system setting - do not modify
Version=10.1.2
```

*Figure S1: Class distribution for the Unspecified Disease cohorts.*

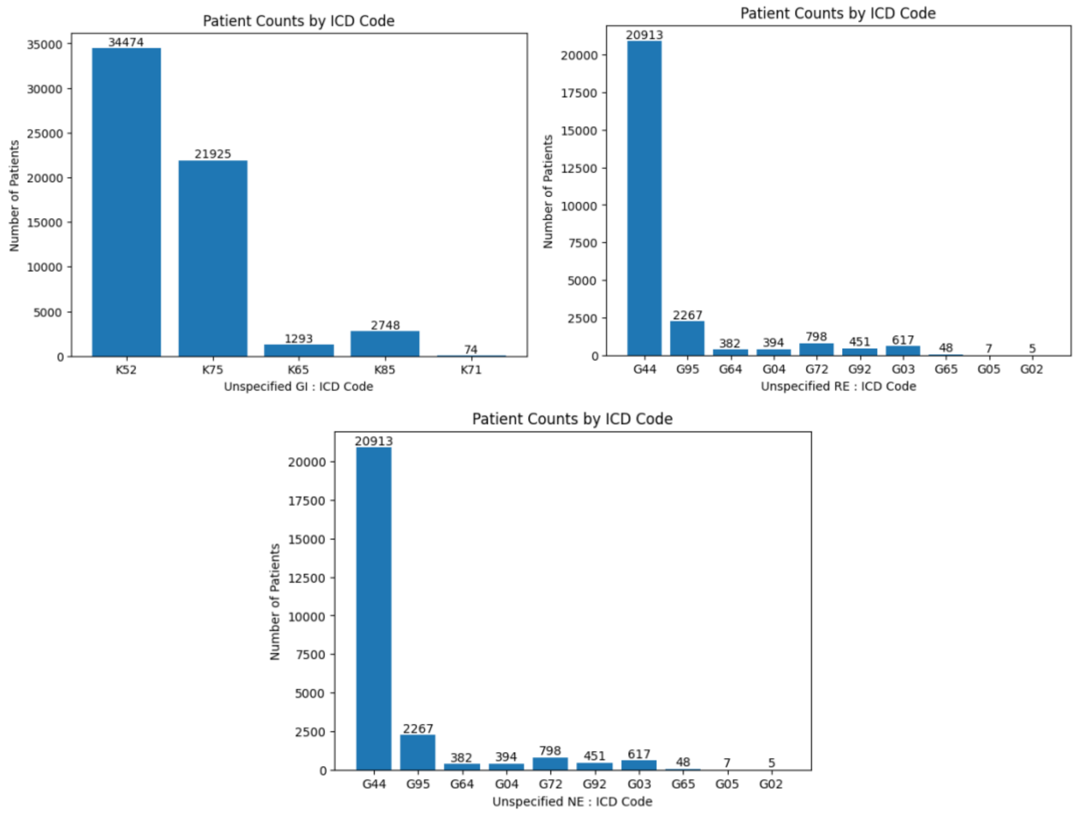

*Figure S2: Class distribution for the Infectious Disease cohort.*

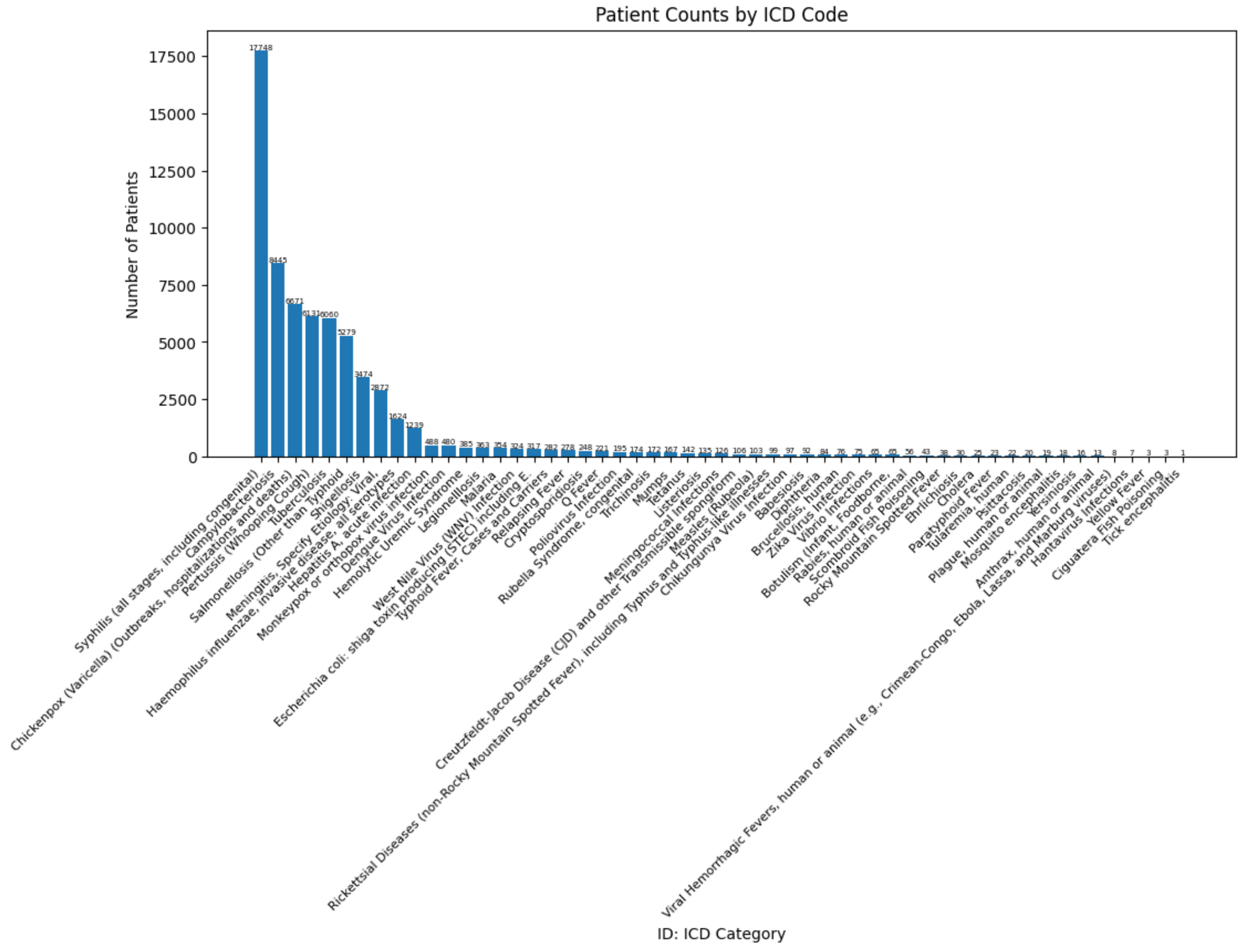
